# Quantized TotalSegmentator for Fast Abdominal Organ Segmentation on CPUs

**DOI:** 10.64898/2026.10.23.26366189

**Authors:** Jeong Hyun Lee

## Abstract

**BACKGROUND:** TotalSegmentator is designed for a graphics processing unit (GPU), runs slowly on a central processing unit (CPU), and depends on Python, limiting CPU-only clinical deployment.

**OBJECTIVE:** The purpose of this study was to evaluate whether TS-Portable, a self-contained CPU executable running 8-bit integer (INT8)–quantized TotalSegmentator models, is noninferior to the reference implementation across 15 abdominal organs in an external multicenter cohort, quantify CPU-only runtime, and demonstrate spleen volumetry on a picture archiving and communication system (PACS) reading station.

**METHODS:** This retrospective study compared TS-Portable with the reference Python implementation on 100 external multicenter CT scans and 28 chronic liver disease series. Per-case mean Dice similarity coefficient (DSC) and 3-mm normalized surface distance (NSD@3 mm) across 15 organs (1.5-mm resolution) and spleen-only mode were tested against a 0.01 noninferiority margin (paired bootstrap 95% CIs). Workstation and PACS runtimes were recorded; spleen volumes were compared via Lin’s concordance correlation coefficient (CCC) and Bland-Altman analysis.

**RESULTS:** The external cohort included 100 patients (53 men, 47 women; mean age, 53 ± 16 [SD] years). Across 15 organs, TS-Portable was noninferior for mean DSC (difference, 0.00017; 95% CI, 0.00003–0.00031; p < .001) and NSD@3 mm (difference, 0.00015; 95% CI, −0.00002 to 0.00031; p < .001) and in spleen-only mode (both p < .001). Workstation 1.5-mm median runtime was 48.0 seconds for TS-Portable on CPU versus 342.5 seconds on CPU (ratio, 7.14) and 61.5 seconds on GPU (ratio, 1.28) for the reference. On PACS, CPU spleen volumetry took 5.8 versus 43.4 seconds, agreeing with the reference (CCC, 0.9999; bias, 0.2%; 95% limits of agreement, −7.2% to 7.6%) and manual contours (CCC, 0.989; bias, −6.8%). In 28 chronic liver disease series, TS-Portable ran from DICOM in 6.5 seconds (median), agreeing with the reference (CCC, 0.99999) and manual contours (CCC, 0.992; bias, −3.4%).

**CONCLUSION:** TS-Portable achieved noninferior abdominal organ segmentation accuracy compared with the reference implementation while running faster on a workstation CPU than the reference on a GPU.

**CLINICAL IMPACT:** Self-contained deployment enables fast multiorgan segmentation on CPU-only computers and spleen volumetry within seconds on a standard reading station, without dedicated GPUs or software installation.

**Highlights:** *Key Finding:* Across 100 external multicenter CT examinations, a quantized, self-contained CPU executable of TotalSegmentator was noninferior to the reference implementation for 15-organ segmentation accuracy and 7.14-fold faster on a workstation CPU, and completed reading-station spleen volumetry in a median of 5.8 seconds.

*Importance:* Compiling and quantizing three-dimensional segmentation models into a standalone CPU executable removes GPU and software-installation barriers, enabling rapid quantitative CT analysis on standard clinical workstations.

## Introduction

Deep learning algorithms based on the nnU-Net framework [1] perform automated multiorgan segmentation on CT examinations with high accuracy, with TotalSegmentator serving as one of the most widely used open-source tools in radiology research [2, 3]. TotalSegmentator is used extensively in research; recently it has been used in the development of advanced artificial intelligence (AI) models, including two recent abdominal CT foundation models. Specifically, Merlin from Stanford was trained and tested for its 20-organ segmentation task on the TotalSegmentator dataset [4], whereas RADAR from Alibaba DAMO Academy used masks produced by the TotalSegmentator tool to pair organ regions with radiological report text during anatomy-aware vision-language pretraining [5].

Since these models generate accurate organ segmentation masks, clinically informative measurements can be derived from them, including organ volume and attenuation [6, 7] as well as radiomic features [8]. For example, spleen volume tracks portal pressure in chronic liver disease and, combined with liver volume, estimates the hepatic venous pressure gradient [9, 10], while a 35% reduction in spleen volume serves as the primary endpoint of Janus kinase inhibitor trials in myelofibrosis [11]. Nevertheless, high computational demands and the complexity of software setup remain major barriers to deploying AI tools in daily clinical practice [12]. The reference implementation is designed for a GPU, runs slowly on a central processing unit (CPU), and relies on a complex software stack that is difficult to install on closed-network hospital computers. Meanwhile, a typical clinical picture archiving and communication system (PACS) reading station contains a standard CPU, often a low-end display GPU, and lacks administrator privileges.

Software engineering techniques can make computationally intensive three-dimensional segmentation models, such as those built on nnU-Net, easier to deploy. The reference implementation of TotalSegmentator is written in Python, a language favored for ease of development rather than speed [13], and runs on the PyTorch deep learning library. A trained model can instead be converted to the Open Neural Network Exchange (ONNX) format, a standard file format for neural networks, and run by a CPU-optimized inference engine [14, 15] within a single program written in Rust, a compiled language [16], so that no Python installation is required. However, to run in practical time on a CPU engine alone, an nnU-Net in a recent challenge had to be retrained at a coarse 2.5-mm resolution [17]. Quantization, in which model weights and activations are stored as lower-fidelity numbers, such as 8-bit integers (INT8) rather than 32-bit floating-point values, further accelerates CPU execution without retraining or lowering resolution, at the cost of numerical rounding. It is under intensive development for large language models [18], but in three-dimensional medical image segmentation its evidence rests largely on simulated rather than deployed inference [19], and the first deployed pipeline, reported only recently, targets only specific GPUs [20]. Because rounding can shift a segmentation boundary, the relevant clinical question is whether segmentation accuracy is preserved, which warrants a noninferiority comparison on external data.

To address this need, we developed TS-Portable, a self-contained CPU executable that runs INT8-quantized versions of the TotalSegmentator models with accuracy controlled during the quantization process. The purpose of this study was to evaluate whether TS-Portable is noninferior to the reference implementation across 15 abdominal organs in an external multicenter cohort, to quantify its runtime and memory consumption on CPU-only computers, and to demonstrate its feasibility for spleen volumetry on a hospital PACS reading station.

## Methods

This retrospective study used only publicly available, deidentified CT datasets. The institutional review board exempted the study from review and waived the requirement for informed consent.

### Study Design

Figure 1 shows the selection of scans and Figure 2 the deployment and study design. TS-Portable runs 8-bit integer (INT8) versions of the 1.5-mm and 3-mm models of TotalSegmentator, version 2.12.0, and was compared with the same models run through the distributed Python package of TotalSegmentator, hereafter the reference implementation. The primary analysis compared segmentations of 15 abdominal organs by the 1.5-mm models in an external multicenter cohort; analyses at 3 mm and in the TotalSegmentator development set were secondary. Once the primary analysis had met the criterion, the spleen was tested in spleen-only mode, the workflow used for spleen volumetry. Runtime and memory were measured on a workstation, and spleen volume agreement and runtime were measured on a picture archiving and communication system (PACS) reading station.

**Fig. 1.**
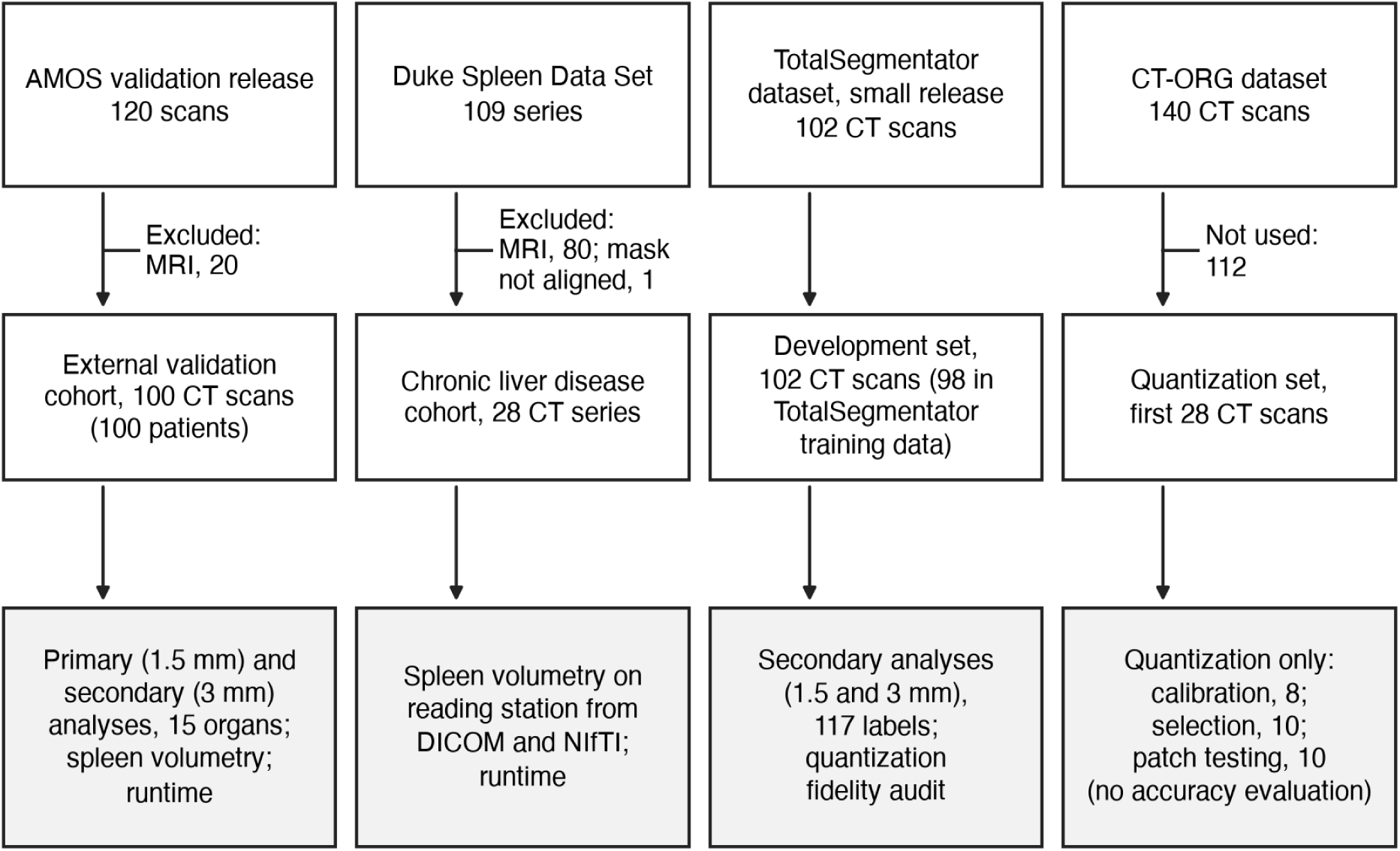
Flowchart shows selection of scans for the external validation cohort, the chronic liver disease cohort, the development set, and the quantization set. AMOS = Abdominal Multi-Organ Segmentation.

**Fig. 2.**
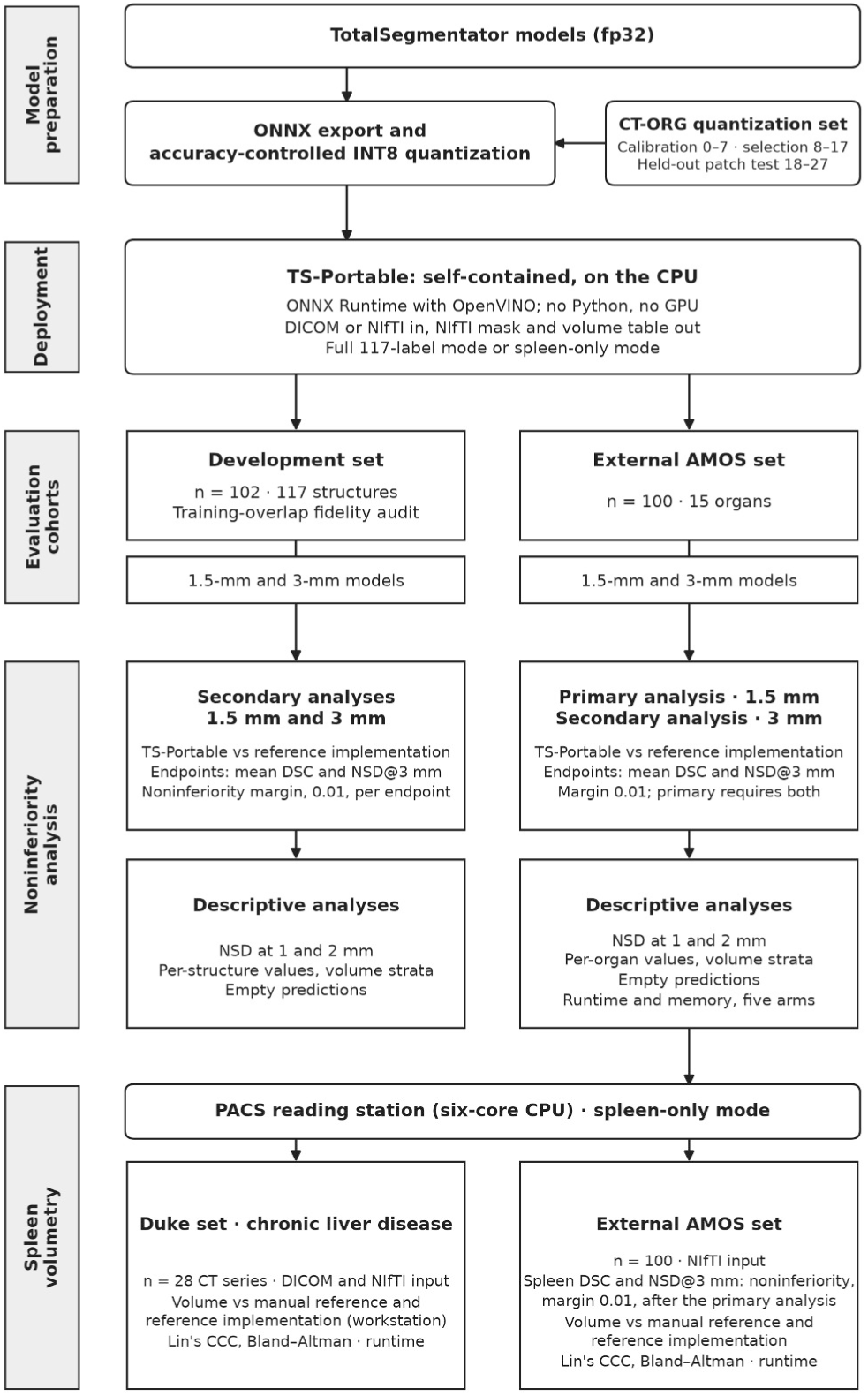
Diagram shows deployment and study design. The fp32 TotalSegmentator models were converted to Open Neural Network Exchange (ONNX) format and quantized with accuracy control on CT-ORG volumes. TS-Portable is the self-contained program that runs the models through ONNX Runtime with OpenVINO on the CPU, reading a DICOM series or NIfTI file and writing a NIfTI mask and a volume table. Both evaluation cohorts were segmented at 1.5 and 3 mm by TS-Portable and by the reference implementation. The primary analysis was the external AMOS analysis at 1.5 mm, with per-case mean DSC and NSD@3 mm as endpoints and a 0.01 margin; once it had met the criterion, the spleen in spleen-only mode was tested with the same margin. Spleen volume agreement and runtime were then measured in spleen-only mode on a PACS reading station, in the AMOS cohort and in 28 CT series of the Duke Spleen Data Set from patients with chronic liver disease; for the Duke series, the reference implementation was run on the workstation. AMOS = Abdominal Multi-Organ Segmentation, CCC = Lin’s concordance correlation coefficient, DSC = Dice similarity coefficient, fp32 = 32-bit floating-point, INT8 = 8-bit integer, NSD@3 mm = normalized surface distance at a 3-mm tolerance, PACS = picture archiving and communication system.

### Datasets

The external validation cohort was the validation release of the Abdominal Multi-Organ Segmentation (AMOS) dataset [21], on which TotalSegmentator was not trained. Of its 120 scans, 20 MRI examinations were excluded, leaving 100 CT scans of 100 patients from two centers. Examination dates were not available in the public metadata (Table S1). Segmentations were evaluated against the AMOS reference masks for the 15 abdominal organs that TotalSegmentator also labels: spleen, right and left kidneys, gallbladder, esophagus, liver, stomach, aorta, inferior vena cava, pancreas, right and left adrenal glands, duodenum, urinary bladder, and prostate. These masks had been produced by model-assisted labeling, checked case by case by five junior radiologists, and validated by three senior radiologists with more than 10 years of experience [21]; they were used unchanged. AMOS uses one label for the prostate and the uterus; a radiologist with 12 years of experience in abdominal CT, blinded to the recorded sex, assigned sex from the pelvic anatomy, and the label was scored as the prostate in men (Supplemental Material, AMOS pelvic label and sex assignment).

The TotalSegmentator development set was the 102-scan small release of the TotalSegmentator dataset [2], evaluated on all 117 labels. Because 98 of the 102 scans were part of TotalSegmentator’s training data, this cohort served as an audit of quantization fidelity rather than as an estimate of accuracy on unseen scans. The first 28 volumes of the CT-ORG dataset [22] were used only to develop the quantized models and were disjoint from both evaluation cohorts.

### TS-Portable

The seven TotalSegmentator models (five 1.5-mm submodels, the 3-mm model, and the 6-mm localization model) were converted to the Open Neural Network Exchange (ONNX) format and are run by a single program written in Rust [16] through ONNX Runtime with OpenVINO (onnxruntime-openvino, version 1.24.1; OpenVINO, version 2025.4.1 [Intel]) [14, 15]. The program is distributed as one folder of about 350 MB, including 212 MB of model files, runs without installation or administrator privileges, reads a DICOM series or NIfTI file, and writes a mask with a per-structure volume table. It reproduces the reference implementation’s processing, including spleen-only mode, in which the 6-mm model localizes the spleen, the scan is cropped around it, and the 1.5-mm model segments the crop.

Quantization stores the model’s weights and intermediate values as 8-bit integers instead of 32-bit floating-point (fp32) values. It was accuracy controlled with the Neural Network Compression Framework (version 3.2.0 [Intel]) [23], which keeps the operations most sensitive to quantization in fp32 (Fig. S1, Table S2). Software versions are listed in Table S3.

### Endpoints

The endpoints were the per-case mean Dice similarity coefficient (DSC) and the per-case mean normalized surface distance at a 3-mm tolerance (NSD@3 mm, also called surface Dice), a surface metric less dependent on structure volume [24]. The tolerance is a property of the metric, distinct from the 1.5-mm and 3-mm model resolutions. Each organ present in the reference mask received equal weight within a case, and an empty prediction scored 0 on both metrics. NSD at 1-mm and 2-mm tolerances, organ-level values, structure-volume strata, direct mask-to-mask agreement between the two implementations, and the largest per-case and per-organ differences were reported descriptively.

### Statistical Analysis

The noninferiority margin was set at 0.01, one percentage point of DSC or NSD, for both endpoints before the analysis. No clinically validated margin exists; 0.01 is a technical threshold chosen conservatively from prior literature [20, 24, 25]. For context, DSC between independent human readers ranges from about 0.6 to 0.97 across these organs [26–31]. The paired difference was the reference implementation minus TS-Portable, so that positive values favor the reference implementation. Paired case bootstrapping with 10,000 resamples gave two-sided 95% bias-corrected and accelerated (BCa) CIs, and one-sided p values against the margin were obtained by inversion of the BCa interval [32, 33]. The primary analysis required a one-sided p < .05 on both endpoints; the three secondary analyses used the same margin, and Holm adjustment was applied across their six endpoint tests. The spleen test was performed only after the primary analysis had met the criterion and therefore needed no adjustment for multiplicity; it paired TS-Portable’s reading-station masks with spleen-only masks from the reference implementation, version 2.12.0, on the workstation. All other organ-level analyses were descriptive. No sample-size calculation was performed; all available scans were analyzed.

In an exploratory analysis, structures were grouped by median reference volume into four strata (cutpoints, 25, 75, and 185 mL) and into hollow organs versus all other structures, and pooled paired differences with BCa intervals were computed for each stratum. NSD was computed with the surface-distance library (version 0.1 [Google DeepMind]), and statistical analyses were performed with R (version 4.1.2) using the boot (version 1.3-28) and boot.pval (version 0.7.0) packages.

### Runtime Benchmarking

Runtime and memory were measured on all 100 AMOS scans on a workstation (Ryzen Threadripper PRO 7945WX 12-core CPU [AMD], 128 GB of RAM, RTX A6000 GPU [NVIDIA]; Pop!_OS 22.04) in five configurations, all with default settings on the same input files: the reference implementation on the GPU and on the CPU; TS-Portable with the unquantized fp32 ONNX models on the CPU and on the GPU; and TS-Portable with its INT8 models on the CPU. The runtime endpoint was the cold end-to-end wall time per case, from process launch to the written mask, including model loading. Memory was the peak proportional set size of the process tree. Runtime was compared as the ratio of each configuration’s median wall time to the median of TS-Portable with INT8 models, with paired case-bootstrap CIs.

### Spleen Volumetry on a PACS Reading Station

TS-Portable was run as a Windows program on a PACS reading station (Core i5-10400 six-core CPU [Intel], 32 GB of RAM, Quadro P400 GPU with 2 GB of memory [NVIDIA]; Windows 11 Enterprise). For the comparison, the reference implementation was run as a self-contained Windows package of the Python program (PyInstaller; 6.4 GB), prepared on another computer. The station ran TotalSegmentator version 2.4.0 because its installed graphics driver could not run the software stack of version 2.12.0; the two versions share identical model weights and gave nearly identical spleen volumes on the workstation (concordance correlation coefficient, 0.99999; Table S4). All 100 AMOS scans were processed from NIfTI files in spleen-only mode and timed for TS-Portable on the CPU and for the reference implementation on the CPU and on the GPU, after one warm-up case. Spleen volume was calculated as the voxel count multiplied by the voxel volume, and the spleen-only masks were also scored by DSC and NSD@3 mm against the manual reference. Agreement between TS-Portable and the manual reference, between the reference implementation and the manual reference, and between the two implementations was assessed with Lin’s concordance correlation coefficient (CCC) with BCa CIs and with the Bland-Altman bias and 95% limits of agreement (LOA), expressed as percentages of the pair mean, with percentile bootstrap CIs from 10,000 case resamples; the 2.5th and 97.5th percentiles of the observed differences were also reported. In sensitivity analyses, agreement was recomputed without the smallest spleen in the cohort, and the spleen noninferiority test was repeated with the reference implementation version 2.4.0 on the reading station as comparator.

Spleen volumetry was also evaluated on the 29 contrast-enhanced axial CT series of the Duke Spleen Data Set [34], drawn from patients with known or suspected chronic liver disease, with or without cirrhosis or portal hypertension; its manual spleen masks were drawn by trained technologists and reviewed by an abdominal radiologist. One series was excluded because its reference mask was not spatially aligned with the image series, leaving 28. On the reading station, TS-Portable processed each series in spleen-only mode from the distributed DICOM files and, with the same timing protocol, from the same series converted to NIfTI. The reference implementation, version 2.12.0, was run in spleen-only mode on the workstation, and agreement was assessed as in the AMOS cohort.

### Code Availability

The TS-Portable program, its source code, and the model weights are available at https://github.com/sigjhl/ts-portable.

## Results

### Study Sample

The external AMOS cohort comprised 100 CT scans of 100 patients (53 men and 47 women; mean age, 53 ± 16 [SD] years; range, 13–94 years) (Table 1). The manual reference spleen volume had a median of 187 mL (IQR, 139–261 mL; range, 16–954 mL). The development set comprised 102 CT scans (63 men, 38 women, and one patient of unspecified sex; mean age, 67 ± 13 [SD] years; range, 23–91 years).

**TABLE 1:** Characteristics of the Evaluation Cohorts.

| Characteristic | External AMOS cohort | Chronic liver disease cohort (Duke) | TotalSegmentator development set |
| --- | --- | --- | --- |
| No. of CT scans (patients) | 100 (100) | 28 (28)<sup>a</sup> | 102 (102) |
| Age (y), mean ± SD (range) | 53 ± 16 (13–94) | Not available | 67 ± 13 (23–91) |
| Sex, men / women | 53 / 47<sup>b</sup> | Not available | 63 / 38<sup>c</sup> |
| Scanner vendor | Siemens Healthineers, 50; Toshiba, 27; Philips, 23 | GE HealthCare, 13; Siemens Healthineers, 13; Philips, 1; Toshiba, 1 | Siemens Healthineers, 100; GE HealthCare, 2 |
| Centers | 2 | 1 | Not available |
| Slice spacing (mm), median (range) | 5.0 (1.25–5.0) | 5.0 (2.5–5.0) | 1.5 (resampled release) |
| In-plane pixel spacing (mm), median (range) | 0.67 (0.45–1.04) | 0.78 (0.63–0.98) | 1.5 (resampled release) |
| Structures evaluated | 15 abdominal organs | Spleen | 117 labels |
| Scans in TotalSegmentator training data | 0 | 0 | 98 |
| Role in this study | Primary and secondary noninferiority analyses; spleen volumetry; runtime | Spleen volumetry from DICOM and NIfTI input; runtime | Secondary noninferiority analyses (quantization fidelity audit) |

Note—Data are numbers of scans unless otherwise indicated. Examination dates were not available in the public metadata; contrast phase was not available for the AMOS and TotalSegmentator cohorts, and the Duke series were post-contrast. AMOS = Abdominal Multi-Organ Segmentation. <sup>a</sup>One of 29 CT series was excluded because its reference mask was not spatially aligned with the image series. The dataset's 69 case identifiers correspond to its 69 patients [34], so each series is from a different patient. <sup>b</sup>AMOS: sex assigned from the pelvic anatomy (Supplemental Material, AMOS pelvic label and sex assignment); it differed from the recorded sex in 27 scans. <sup>c</sup>Sex was unspecified for one patient.

### Noninferiority

All four analyses met the noninferiority criterion on both endpoints (Table 2), and all six secondary endpoint tests remained significant after Holm adjustment (adjusted p ≤ .003). In the primary analysis, the per-case mean DSC was 0.8493 for TS-Portable and 0.8495 for the reference implementation (difference, 0.00017; 95% CI, 0.00003–0.00031; p < .001), and the mean NSD@3 mm was 0.9086 and 0.9088, respectively (difference, 0.00015; 95% CI, −0.00002 to 0.00031; p < .001). The largest one-sided upper bound across the four analyses was 0.00069. For the spleen in spleen-only mode, the DSC was 0.9441 for TS-Portable and 0.9433 for the reference implementation (difference, −0.00081; 95% CI, −0.00567 to 0.00054), and the NSD@3 mm was 0.9615 and 0.9616 (difference, 0.00009; 95% CI, −0.00294 to 0.00150); both met the margin (one-sided upper bounds, 0.00045 and 0.00131; p < .001). The unquantized fp32 ONNX models run by the same program differed from the reference implementation by at most 0.00012 (Table S5). Figure 3 shows the spleen and liver of one patient from the AMOS cohort, and Figure S2 shows further examples at 1.5 and 3 mm.

**Fig. 3.**
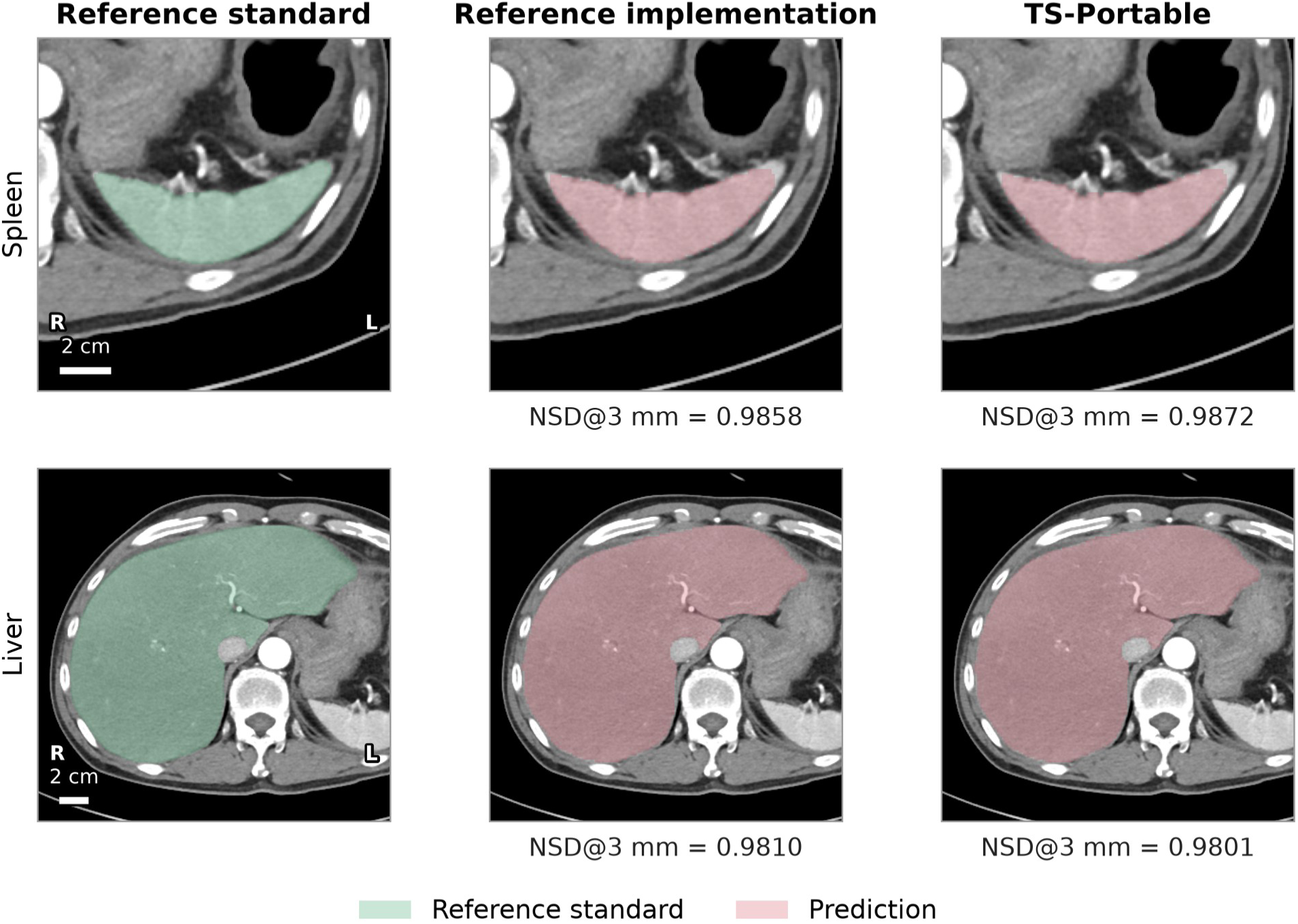
Man in his 60s from external AMOS cohort. Axial CT images show spleen (top) and liver (bottom) segmentation at 1.5 mm in full 117-label mode: manual reference standard (green, left), reference implementation (pink, center), and TS-Portable (pink, right). Case was selected as closest to cohort median spleen NSD@3 mm; each organ is shown at its largest axial cross-section. NSD@3 mm values, computed on whole three-dimensional masks, are shown below segmentations. Further examples are in Figure S2. AMOS = Abdominal Multi-Organ Segmentation, NSD@3 mm = normalized surface distance at a 3-mm tolerance.

**TABLE 2:** Segmentation Accuracy Against the Reference Masks and Paired Noninferiority Comparison of TS-Portable With the Reference Implementation.

| Analysis | n | Endpoint | Reference implementation | TS-Portable | Difference (95% CI)<sup>a</sup> | One-sided 95% upper bound<sup>b</sup> |
| --- | --- | --- | --- | --- | --- | --- |
| **All structures, per-case mean** |  |  |  |  |  |  |
| External AMOS, 1.5 mm<sup>c</sup> | 100 | DSC | 0.8495 | 0.8493 | 0.00017 (0.00003–0.00031) | 0.00029 |
| External AMOS, 1.5 mm<sup>c</sup> | 100 | NSD@3 mm | 0.9088 | 0.9086 | 0.00015 (–0.00002 to 0.00031) | 0.00028 |
| External AMOS, 3 mm | 100 | DSC | 0.8097 | 0.8095 | 0.00011 (–0.00033 to 0.00059) | 0.00051 |
| External AMOS, 3 mm | 100 | NSD@3 mm | 0.8856 | 0.8856 | –0.00004 (–0.00063 to 0.00057) | 0.00047 |
| Development set, 1.5 mm | 102 | DSC | 0.9146 | 0.9145 | 0.00010 (0.00002–0.00019) | 0.00017 |
| Development set, 1.5 mm | 102 | NSD@3 mm | 0.9550 | 0.9550 | 0.00003 (–0.00003 to 0.00011) | 0.00010 |
| Development set, 3 mm | 102 | DSC | 0.8289 | 0.8285 | 0.00041 (0.00030–0.00066) | 0.00061 |
| Development set, 3 mm | 102 | NSD@3 mm | 0.9571 | 0.9568 | 0.00028 (0.00013–0.00086) | 0.00069 |
| **Spleen, spleen-only mode, external AMOS<sup>d</sup>** |  |  |  |  |  |  |
| External AMOS, 1.5 mm | 100 | DSC | 0.9433 | 0.9441 | –0.00081 (–0.00567 to 0.00054) | 0.00045 |
| External AMOS, 1.5 mm | 100 | NSD@3 mm | 0.9616 | 0.9615 | 0.00009 (–0.00294 to 0.00150) | 0.00131 |

Note—Per-case means are over 15 organs (AMOS) or 117 labels (development set). DSC and NSD@3 mm are computed against the dataset reference masks; direct mask-to-mask agreement between implementations is in Table S6. AMOS = Abdominal Multi-Organ Segmentation, DSC = Dice similarity coefficient, NSD@3 mm = normalized surface distance at a 3-mm tolerance. <sup>a</sup>Reference implementation minus TS-Portable; positive values favor the reference implementation. Two-sided 95% bias-corrected and accelerated bootstrap CI from 10,000 paired case resamples. <sup>b</sup>Upper bound of the one-sided 95% CI, compared with the 0.01 noninferiority margin; the one-sided p value against the margin was < .001 in every row. <sup>c</sup>Primary analysis; noninferiority required both endpoints to meet the margin. <sup>d</sup>Tested after the primary analysis with the same procedure, margin, and criterion. In the sensitivity analysis with reference
implementation version 2.4.0 on the reading station as comparator, the upper bounds were 0.00027 and 0.00092.

An empty prediction was an organ that was present in the reference mask but had no voxels in the predicted mask. In the AMOS cohort, both implementations had empty predictions in the same organ-case pairs: 1 of 1440 pairs at 1.5 mm (a gallbladder) and 7 of 1440 pairs at 3 mm (gallbladder, 6; left adrenal gland, 1). In the development set (9299 pairs), TS-Portable had two more empty predictions than the reference implementation at 1.5 mm (69 vs 67) and one more at 3 mm (112 vs 111). All empty predictions of the reference implementation were also empty with TS-Portable.

### Agreement Between Implementations

Compared directly, mask to mask, the two implementations had a mean DSC of 0.996 over the 15 AMOS organs at 1.5 mm and of 0.996 over the 117 development-set labels (Table S6). In the primary analysis, the per-case difference was at most 0.0023 for DSC and 0.0025 for NSD@3 mm. At 3 mm, the per-case difference in NSD@3 mm was more than 0.01 in one AMOS case and in one development-set case (Table S7). For individual organs in single cases of the primary analysis, the largest loss with TS-Portable was 0.028 for DSC (a gallbladder) and 0.041 for NSD@3 mm (a right adrenal gland). The largest loss in all analyses was in a C6 vertebra at 3 mm in the development set: TS-Portable left it empty, and the reference implementation had a DSC of 0.44.

### Descriptive Accuracy Analyses

At NSD tolerances of 1 mm and 2 mm, differences were at most 0.0006, with upper limits of at most 0.0009 (Table S8). The largest organ-level DSC difference in the AMOS cohort at 1.5 mm was for the inferior vena cava (difference, 0.0032; 95% CI, 0.0027–0.0038; Tables S9 and S10). Across structure-volume strata and the hollow-organ grouping, every pooled difference had an upper bound of at most 0.0013 (Fig. S3, Table S11). One structure, the left 12th rib in the development set, had an NSD@3 mm upper limit above the margin (difference, 0.0021; 95% CI, 0.0002–0.0109), attributable to one scan in which both implementations misassigned rib numbers (Fig. S4).

### Runtime and Memory on the Workstation

At 1.5 mm, the median wall time was 48.0 seconds for TS-Portable on the CPU, 342.5 seconds for the reference implementation on the CPU (ratio of medians, 7.14; 95% CI, 7.08–7.21), and 61.5 seconds for the reference implementation on the GPU (ratio, 1.28; 95% CI, 1.25–1.35) (Table 3, Fig. 4). At 3 mm, the corresponding medians were 3.9, 27.3, and 16.1 seconds. Of the sevenfold gain on the CPU at 1.5 mm, the program and its inference library accounted for a 3.5-fold gain (fp32 ONNX models on the CPU, 98.0 seconds) and quantization for a further 2.0-fold gain; at 3 mm, the gains were 4.3-fold and 1.6-fold. The median peak memory was 3.9 GiB for TS-Portable at both resolutions, compared with 7.1 GiB at 1.5 mm and 6.3 GiB at 3 mm for the reference implementation on the CPU.

**Fig. 4.**
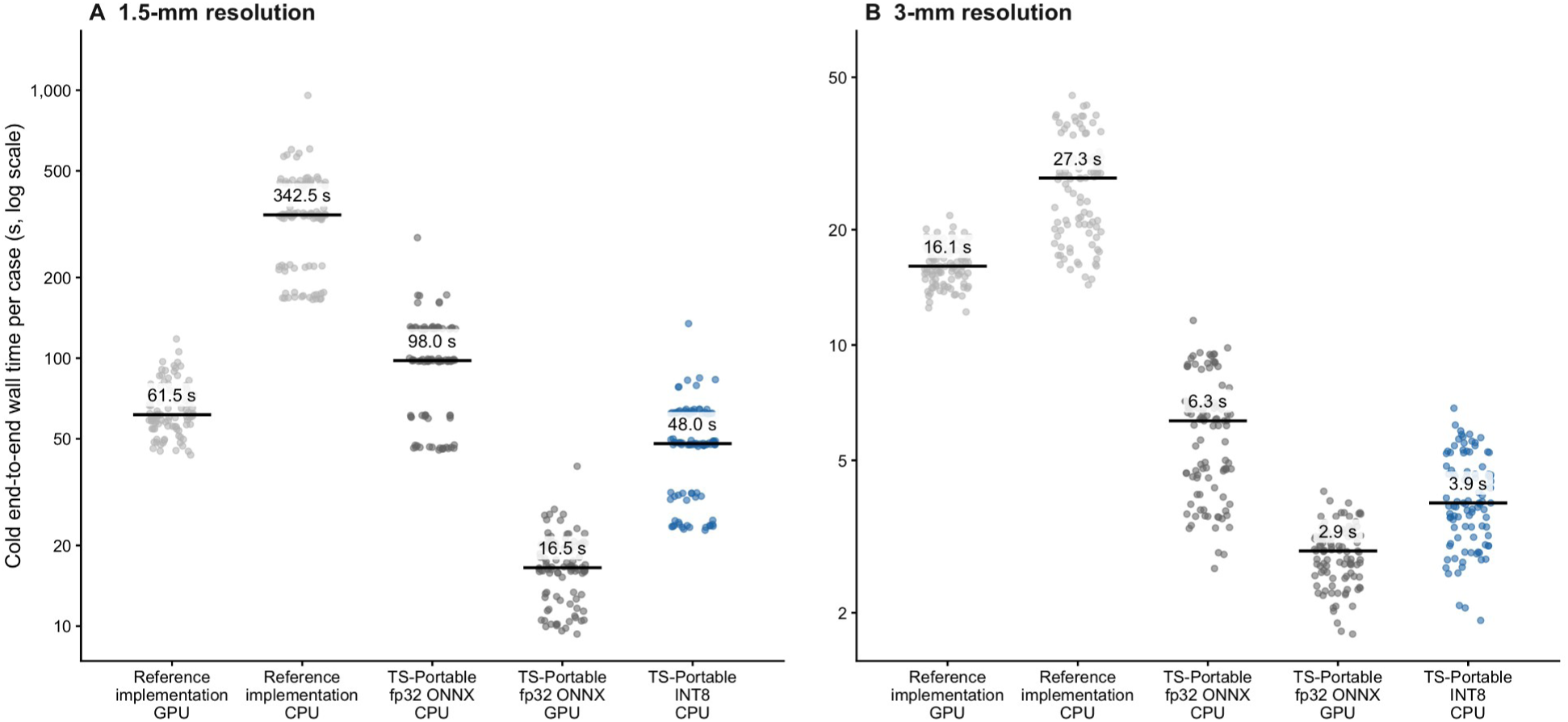
Dot plots show per-case cold end-to-end wall time for five configurations on all 100 external AMOS scans (logarithmic axis). Each point is one scan (gray, comparator configurations; blue, TS-Portable with INT8 models); horizontal bars mark medians, whose values are printed above them. AMOS = Abdominal Multi-Organ Segmentation, fp32 = 32-bit floating-point, INT8 = 8-bit integer, ONNX = Open Neural Network Exchange. A, Results at 1.5 mm. B, Results at 3 mm.

**TABLE 3:** Cold End-to-End Runtime and Memory on the Workstation, 100 External AMOS Scans.

| Configuration | Inference library | Device | 1.5 mm, median wall time (s) | 1.5 mm, ratio (95% CI)<sup>a</sup> | 3 mm, median wall time (s) | 3 mm, ratio (95% CI)<sup>a</sup> | Peak memory, 1.5 mm / 3 mm (GiB) |
| --- | --- | --- | --- | --- | --- | --- | --- |
| Reference implementation | PyTorch | GPU | 61.5 | 1.28 (1.25–1.35) | 16.1 | 4.15 (3.81–4.36) | 7.8 / 5.6<sup>b</sup> |
| Reference implementation | PyTorch | CPU | 342.5 | 7.14 (7.08–7.21) | 27.3 | 7.05 (6.58–7.25) | 7.1 / 6.3 |
| TS-Portable, fp32 ONNX | ONNX Runtime with OpenVINO | CPU | 98.0 | 2.04 (2.03–2.05) | 6.3 | 1.64 (1.60–1.72)<sup>c</sup> | 4.6 / 4.3 |
| TS-Portable, fp32 ONNX | ONNX Runtime with CUDA | GPU | 16.5 | 0.34 (0.34–0.35) | 2.9 | 0.75 (0.71–0.79) | 3.4 / 2.6<sup>b</sup> |
| TS-Portable, INT8 | ONNX Runtime with OpenVINO | CPU | 48.0 | 1 (reference) | 3.9 | 1 (reference) | 3.9 / 3.9 |

Note—Wall time is from process launch through writing of the output mask, one case per process; each configuration processed all 100 scans once. Workstation: Ryzen Threadripper PRO 7945WX 12-core CPU (AMD), 128 GB of RAM, RTX A6000 GPU (NVIDIA). Memory is the median per-case peak proportional set size of the process tree. AMOS = Abdominal Multi-Organ Segmentation, CUDA = GPU execution path of ONNX Runtime, fp32 = 32-bit floating-point, INT8 = 8-bit integer, ONNX = Open Neural Network Exchange. <sup>a</sup>Ratio of the configuration's median to the median of TS-Portable with INT8 models on the CPU; values above 1 favor TS-Portable with INT8 models. Two-sided 95% bias-corrected and accelerated CIs from 10,000 paired case resamples. <sup>b</sup>Host process memory only; GPU memory is not included. <sup>c</sup>Runtimes at 3 mm cluster by input volume, so this interval is approximate.

### Spleen Volumetry on the PACS Reading Station

In spleen-only mode on the reading station, the median runtime per case was 5.8 seconds for TS-Portable on the CPU (90th percentile, 7.0 seconds), 43.4 seconds for the reference implementation on the CPU, and 43.5 seconds for the reference implementation on the Quadro P400 GPU, which it used without falling back to the CPU, although the GPU’s 2 GB of memory is below the reference implementation’s stated requirement [3] (Table 4).

**TABLE 4:** Spleen-Only Mode Runtime and Memory on the PACS Reading Station, 100 External AMOS Scans.

| Configuration | Device | Median wall time (s) | 90th percentile (s) | Ratio of medians to TS-Portable | Median peak working set (GiB) |
| --- | --- | --- | --- | --- | --- |
| TS-Portable, INT8 | CPU | 5.8 | 7.0 | 1 (reference) | 1.46 |
| Reference implementation, version 2.4.0 | CPU | 43.4 | 47.1 | 7.49 | 4.11 |
| Reference implementation, version 2.4.0 | GPU | 43.5 | 46.3 | 7.50 | 4.78<sup>a</sup> |

Note—Reading station: Core i5-10400 six-core CPU (Intel), 32 GB of RAM, Quadro P400 GPU with 2 GB of memory (NVIDIA), Windows 11 Enterprise. One warm-up case was run, then the 100 cases in sequence, one case per process. Windows working-set memory is not comparable with the proportional set size in Table 3. AMOS = Abdominal Multi-Organ Segmentation, INT8 = 8-bit integer, PACS = picture archiving and communication system. <sup>a</sup>Peak GPU memory was 1.93 GiB.

Spleen volumes from TS-Portable and the reference implementation agreed on the station with a CCC of 0.9999 (95% CI, 0.9998–1.0000), a relative bias of 0.2%, and 95% LOA of −7.2% to 7.6% (Table 5, Fig. 5). Against the manual reference, the CCC was 0.989 (95% CI, 0.978–0.994) for both TS-Portable and the reference implementation, with relative biases of −6.8% for both and 95% LOA of −28.5% to 15.0% and −32.8% to 19.1%, respectively. Because the differences were skewed, the 2.5th and 97.5th percentiles of the observed differences were also calculated: −20.6% to 0.9% for TS-Portable and −20.6% to 1.0% for the reference implementation against the manual reference, and −1.4% to 1.5% between implementations. The median absolute difference between TS-Portable and the manual reference was 8.9 mL (5.0%), and 13 of 100 volumes differed by more than 10%. Excluding the smallest spleen (16 mL by manual reference; 4.9 mL by TS-Portable and 3.4 mL by the reference implementation), the 95% LOA against the manual reference were −15.1% to 3.6% for TS-Portable and −14.7% to 3.5% for the reference implementation, and −2.5% to 2.2% between implementations. Both implementations segmented this spleen correctly in full 117-label mode but undersegmented it in spleen-only mode, although the crop contained the entire spleen (Fig. S5). In full mode on the workstation, the 95% LOA against the manual reference were approximately −17% to 5% for both implementations (Table S12).

**Fig. 5.**
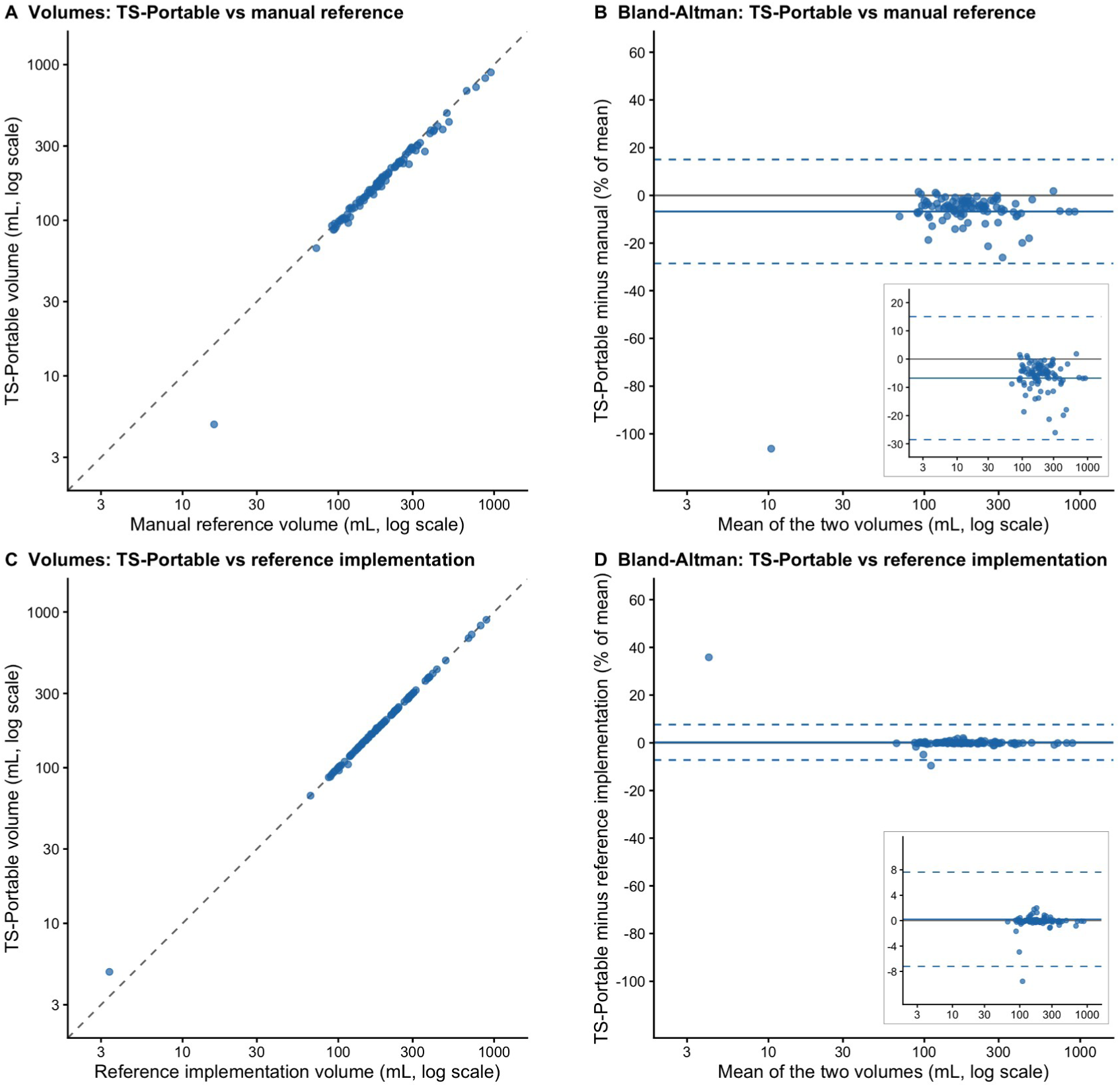
Spleen volumetry in spleen-only mode on PACS reading station, 100 external AMOS scans. Volumes are shown on logarithmic axes with dashed identity line (A and C); Bland-Altman plots show difference as percentage of pair mean against pair mean on logarithmic axis, with zero (gray), mean difference (solid blue), and 95% limits of agreement (dashed blue) (B and D). Isolated point in each panel is smallest spleen in cohort (16 mL by manual reference; 4.9 mL by TS-Portable and 3.4 mL by reference implementation). Insets in B and D repeat plot with vertical axis restricted to central distribution and omit only that case; lines are those of all 100 scans. AMOS = Abdominal Multi-Organ Segmentation, PACS = picture archiving and communication system. A, Volume of TS-Portable against manual reference volume. B, Bland-Altman plot of TS-Portable against manual reference. C, Volume of TS-Portable against volume from reference implementation run on same station. D, Bland-Altman plot of TS-Portable against reference implementation run on same station.

**TABLE 5:** Spleen Volume Agreement in Spleen-Only Mode on the PACS Reading Station.

| Comparison | CCC (95% CI)<sup>a</sup> | Relative bias, % (95% CI)<sup>b</sup> | Lower 95% LOA, % (95% CI)<sup>b</sup> | Upper 95% LOA, % (95% CI)<sup>b</sup> | Empirical 2.5th to 97.5th percentile, %<sup>c</sup> | Median absolute difference, mL (% of pair mean) |
| --- | --- | --- | --- | --- | --- | --- |
| **External AMOS cohort (n = 100)** |  |  |  |  |  |  |
| TS-Portable vs manual reference | 0.989 (0.978–0.994) | –6.8 (–9.2 to –5.1) | –28.5 (–44.4 to –12.9) | 15.0 (2.2–26.7) | –20.6 to 0.9 | 8.9 (5.0) |
| Reference implementation vs manual reference | 0.989 (0.978–0.994) | –6.8 (–9.8 to –4.9) | –32.8 (–52.6 to –12.5) | 19.1 (2.1–33.8) | –20.6 to 1.0 | 9.1 (4.9) |
| TS-Portable vs reference implementation | 0.9999 (0.9998–1.0000) | 0.2 (–0.3 to 1.1) | –7.2 (–11.7 to –1.0) | 7.6 (1.0–13.3) | –1.4 to 1.5 | 0.3 (0.2) |
| **Duke cohort, chronic liver disease (n = 28)<sup>d</sup>** |  |  |  |  |  |  |
| TS-Portable vs manual reference | 0.992 (0.986–0.997) | –3.4 (–4.9 to –2.0) | –11.4 (–14.1 to –7.8) | 4.6 (2.5–6.2) | –12.1 to 2.5 | 13.3 (3.1) |
| Reference implementation vs manual reference | 0.992 (0.987–0.997) | –3.2 (–4.7 to –1.8) | –11.1 (–14.0 to –7.4) | 4.7 (2.5–6.3) | –12.1 to 2.6 | 12.6 (3.3) |
| TS-Portable vs reference implementation | 0.99999 (0.99997–0.99999) | –0.2 (–0.4 to –0.1) | –1.1 (–1.8 to –0.3) | 0.7 (0.1–1.1) | –1.2 to 0.1 | 0.6 (0.1) |

Note—Relative bias and 95% LOA (mean ± 1.96 SD) are the first-named minus the second-named volume, as a percentage of the pair mean. In the AMOS cohort, the reference implementation was version 2.4.0 on the reading station. Agreement in full 117-label mode is in Table S12. AMOS = Abdominal Multi-Organ Segmentation, CCC = Lin's concordance correlation coefficient, LOA = limits of agreement, PACS = picture archiving and communication system. <sup>a</sup>Bias-corrected and accelerated bootstrap CI. <sup>b</sup>Percentile bootstrap CI from 10,000 case resamples. <sup>c</sup>Percentiles of the observed relative differences, without a normality assumption. With 28 series in the Duke cohort, they lie close to the smallest and largest differences. <sup>d</sup>TS-Portable from DICOM input on the reading station; reference implementation version 2.12.0 on the workstation.

### Spleen Volumetry in Chronic Liver Disease

In the 28 Duke CT series, manual spleen volumes ranged from 175 to 1598 mL (median, 581 mL), and six exceeded 1000 mL. TS-Portable agreed with the reference implementation with a CCC of 0.99999 (95% CI, 0.99997–0.99999) and 95% LOA of −1.1% to 0.7% (Table 5). Against the manual reference, the CCC was 0.992 (95% CI, 0.986–0.997) for TS-Portable and 0.992 (95% CI, 0.987–0.997) for the reference implementation, with relative biases of −3.4% and −3.2% and 95% LOA of −11.4% to 4.6% and −11.1% to 4.7%, respectively. The masks from DICOM and NIfTI input were identical in every series, and the median runtime from DICOM was 6.5 seconds (IQR, 4.9–8.6 seconds), 0.1 seconds longer than from NIfTI (Table S13).

## Discussion

In this external multicenter study, TS-Portable, a quantized, self-contained CPU deployment of TotalSegmentator, achieved noninferior segmentation accuracy across 15 abdominal organs compared with the reference implementation while substantially reducing execution time and memory consumption on CPU-only hardware. Furthermore, the standalone executable ran faster on a workstation CPU than the reference implementation on a dedicated GPU and measured spleen volume within seconds, in close agreement with the reference implementation, directly on a standard clinical PACS reading station without requiring a Python environment or software installation.

The observed acceleration derived from two separable factors. The compiled inference stack and standalone executable accounted for a 3.5-fold speedup at 1.5-mm resolution, while INT8 quantization contributed an additional 2.0-fold gain. Unquantized models executed within the same standalone program differed from the reference implementation by at most 0.00012, indicating that most of the small accuracy difference at 1.5 mm is attributable to quantization; in the AMOS cohort at 3 mm, the conversion and quantization differences in DSC were of similar size.

These findings address a methodological gap identified in a recent systematic review of quantization in medical imaging, which noted that computational efficiency is frequently estimated through simulation and accuracy is often reported without confidence intervals or external validation [19]. In the present study, the quantized models were evaluated as a deployed executable on external multicenter data, using bootstrap confidence intervals tested against a prespecified noninferiority margin. Although no clinically validated noninferiority margin exists for segmentation agreement and the 0.01 threshold was selected as a conservative technical boundary based on prior literature [20, 24, 25], the empirical differences in our study were an order of magnitude smaller, establishing noninferiority of the per-case mean endpoints far below this margin. Even at the organ level in the external cohort at 1.5 mm, the largest DSC discrepancy occurred in the inferior vena cava, with a mean difference of only 0.003.

Beyond mean differences, the secondary descriptive analyses identified rare case-level divergences. In the development dataset, the left 12th rib exhibited a surface-distance upper confidence limit exceeding the 0.01 margin, driven by a single examination in which both implementations misassigned rib numbering. Empty predictions occurred in the exact same organ-case pairs for both implementations in the external cohort, demonstrating that quantization introduced no new empty predictions there; in the development dataset, quantization resulted in two additional empty organ-case pairs at 1.5-mm resolution out of more than 9,000 total pairs. These rare failures in single cases underscore the importance of reporting distribution tails in addition to mean values when validating deployed models.

In spleen volumetry, TS-Portable showed the same bias and failure modes as the reference implementation against manual contours in both cohorts, indicating that the underestimation of spleen volume comes from the underlying TotalSegmentator model or the annotation convention rather than quantization. The AMOS limits of agreement were wider than the ±4% reported for dedicated deep-learning spleen volumetry [35], partly because both implementations undersegmented the same 16-mL spleen; without it, the limits were still −15.1% to 3.6%, reflecting the shared systematic underestimation. Because TS-Portable carries over the errors of the reference implementation, an implausible volume in practice calls for inspection of the mask on the source images.

More broadly, an offline executable of this design reduces reliance on complex software integration and local computing infrastructure [12] and may widen access to quantitative imaging where dedicated GPUs are lacking [36]. It could also serve radiology AI agents. Because general-purpose vision-language models remain unreliable readers of radiological images [37, 38], one research direction is for agents to call specialized image models as tools [39, 40], and a CPU executable could supply organ masks and volumes where GPUs are scarce. Finally, running this and other three-dimensional imaging models on hospital computers would let research studies export measurements instead of images, which carry a residual risk of reidentification [41] and are far larger to transfer.

This study has several limitations. The prespecified noninferiority margin was a technical threshold rather than a clinically validated boundary, and aggregate differences below this margin do not preclude larger discrepancies in individual organs or cases, which we consequently reported in detail. External validation encompassed 15 abdominal organs; agreement across the remaining 102 anatomic labels was evaluated on the development dataset, the majority of which was included in TotalSegmentator’s training data, and thus represents an audit of quantization fidelity rather than segmentation accuracy on unseen examinations. Spleen volumetry was validated against research contours, and the chronic liver disease cohort was small, originated from a single institution, and included only one spleen above 1500 mL; furthermore, hematologic malignancy and post-intervention spleens were not represented, and low-dose and motion-degraded examinations were not separately evaluated. The reading-station evaluation was performed on a single reading station at one institution, using an older TotalSegmentator version necessitated by the local graphics driver, and the reading station’s CPU lacks the dedicated INT8 instruction sets of newer processors, which may execute the quantized models even more rapidly. Finally, reader performance, clinical workflow integration, longitudinal volumetric tracking, and patient outcomes were not assessed, and TS-Portable remains a research implementation rather than a regulatory-cleared medical device.

In conclusion, a quantized, self-contained CPU deployment of TotalSegmentator achieved noninferior segmentation accuracy compared with the reference implementation across abdominal organs on external multicenter CT examinations and executed faster on a workstation CPU than the reference implementation on a dedicated GPU; on a conventional clinical reading station, it measured spleen volume within seconds.

## Declaration of Generative AI Use

During the preparation of this work the author used Claude (Claude Opus 5.5 and Claude Fable 5.1; Anthropic, San Francisco, CA), Codex (GPT-6-Astra; OpenAI, San Francisco, CA), and Gemini (Gemini 4 Argon; Google, Mountain View, CA) for software development, analysis scripting, language editing, and document preparation. The tools were not used to generate study data, statistical results, or conclusions. The author reviewed and edited all content and takes full responsibility for the manuscript.

## Author contributions

The sole author meets the ICMJE criteria for authorship and performed all parts of the study.

## Data availability

All CT data are public: the Abdominal Multi-Organ Segmentation (AMOS) validation set (Zenodo 7262581), the TotalSegmentator dataset small release (Zenodo 10047263), CT-ORG (The Cancer Imaging Archive, https://doi.org/10.7937/tcia.2019.tt7f4v7o), and the Duke Spleen Data Set (Zenodo 7636640, available on request under its data use terms). The TS-Portable program, its source code, and the ONNX and 8-bit integer model weights are openly available at https://github.com/sigjhl/ts-portable.

## Acknowledgments

The author thanks Jakob Wasserthal and colleagues for releasing TotalSegmentator and its dataset, and the authors of the AMOS, CT-ORG, and Duke Spleen Data Set datasets for making their data publicly available.

