## Supplementary Material for "Quantized TotalSegmentator for Fast Abdominal Organ Segmentation on CPUs"

### Supplemental File for Review

#### Accuracy-controlled quantization

Quantization of the seven models (five 1.5-mm submodels, the 3-mm model, and the 6-mm localization model) used NNCF 3.2.0 `quantize_with_accuracy_control` with ONNX 1.22.0, ONNX Runtime 1.27.0, and Python 3.12.11; deployed inference used `onnxruntime-openvino` 1.24.1 with OpenVINO 2025.4.1 (Supplemental Table S3).

Settings were `QuantizationPreset.MIXED`, `TargetDevice.CPU`, fast bias correction, a maximum accuracy drop of 0.005, and at most four restoration iterations; the last two values were chosen for this study. The first encoder convolution and the final segmentation convolution of each model were never quantized. The final graphs had per-channel INT8 weights and UINT8 activations; all other operations stayed in fp32. Supplemental Table S2 counts the convolutional layers kept in fp32 per model.

The NNCF loop ran on mainline ONNX Runtime because its candidate graphs use signed INT8 activation zero points, which OpenVINO's CPU plugin cannot run. The final models were re-encoded with UINT8 zero points by shifting each activation zero point by 128 with scales unchanged, which represents the same quantized values, re-checked under `onnxruntime-openvino` 1.24.1 on the selection and held-out patches, and released. Four models reached the 0.005 target and three (the 6-mm, 3-mm, and 1.5-mm organ models) stopped at the four-iteration limit (selection-patch fidelity on the quantization runtime, 0.984 to 0.991); on the deployed runtime, selection-patch fidelity was 0.984 to 0.997 and held-out patch fidelity 0.948 to 0.987, and the achieved fidelities on both runtimes are in Supplemental Table S2B. Patch fidelity scores every label fragment in a small patch and served only for model selection. Across the seven models, 154 of 186 convolutional layers were quantized; the remainder were the first and last layer of each model, never quantized, and 18 layers restored by accuracy control (Supplemental Table S2). Every TS-Portable INT8 accuracy and runtime value in the manuscript comes from these released files under that runtime.

#### CT-ORG calibration, selection, and patch testing

The CT-ORG split was fixed before the final models were generated. Volumes 0–7 gave calibration patches, volumes 8–17 gave accuracy-control selection patches, and volumes 18–27 gave held-out test patches that NNCF never saw. These 28 volumes were enough for patch-level calibration, and no more were used; CT-ORG supports no accuracy claim. Calibration used 160 foreground patches per 1.5-mm model, 66 for the 3-mm model, and 44 for the 6-mm model. Selection and patch testing each used two foreground-rich patches per volume, 20 per model. A patch's foreground fraction was the share of its voxels above the patch's minimum normalized intensity (tissue rather than air or padding); patches with more than 3% were ranked by it, and no CT-ORG labels were used.

The accuracy-control objective was the mean foreground Dice between candidate and fp32 patch predictions, averaged over the selection patches, with background excluded. It measured how faithfully a candidate reproduced the fp32 model; it is not a clinical endpoint, not a whole-volume estimate, and not a measure of accuracy against anatomy. The CT-ORG volumes were disjoint from the AMOS and TotalSegmentator evaluation cohorts.

#### INT8 and fp32 execution and precision

The INT8 experiments forced fp32 inference precision for all non-quantized operations. Primitive-level tracing showed UINT8/SINT8 quantized convolutions and fp32 for the remaining convolutions and deconvolutions; no 16-bit floating-point primitive appeared. Resampling, sliding-window tiling, postprocessing, and axis alignment were the same for the fp32 and INT8 pathways. Supplemental Table S5 shows how closely the fp32 ONNX pathway matches the reference implementation; the differences in the main text at 1.5 mm are therefore mostly attributable to quantization rather than to the conversion or the executable; in the AMOS cohort at 3 mm, the conversion and quantization differences in DSC were of similar size.

#### AMOS pelvic label and sex assignment

AMOS uses a single label for the prostate and the uterus. A radiologist with 12 years of experience in abdominal CT reviewed this label on all 100 AMOS CT scans, on contiguous axial slices of the pelvis with the label outlined and sagittal and coronal reformations, blinded to the recorded sex, the site, and the segmentation outputs. Sex was assigned from the pelvic anatomy in the 99 scans that covered the pelvis; the remaining scan ended above the pelvis, had no pelvic label, kept its recorded sex (male), and is the one man without a labeled prostate. The assignment differed from the sex recorded in the AMOS metadata in 27 scans (18 recorded as men, 9 as women), 25 of them from one of the two AMOS sites. The assigned sex was used throughout the study, including Table 1, and the label was scored as the prostate in the 52 men with a labeled prostate. Four women had undergone hysterectomy; in two of them the label covered the vagina (13 and 20 mL) and, as in all women, was not scored as a prostate. With the recorded sex instead, the primary differences were 0.00015 (95% CI, 0.00001–0.00029) for DSC and 0.00013 (95% CI, −0.00003 to 0.00029) for NSD@3 mm, with one-sided upper bounds of 0.00027 for both.

Structures absent from the AMOS reference mask are not scored. With sex assigned as above, 13 organ–case pairs had no reference structure (gallbladder 7, right kidney 2, stomach, duodenum, urinary bladder, and prostate 1 each). TS-Portable or the reference implementation predicted the structure in 7 of them. On review by the same radiologist, six predictions did not correspond to the organ (gallbladder 4, stomach 1, right kidney 1; 0.01–120 mL). In one scan after distal gastrectomy the duodenum was present but unlabeled and difficult to delineate; it was treated as absent. In each of these 7 pairs, both implementations predicted the structure, except for one 0.01-mL gallbladder predicted by the reference implementation only.

#### Paired case bootstrap noninferiority analysis

For each case, the paired difference was reference implementation minus TS-Portable in per-case mean DSC and in per-case mean NSD@3 mm; positive values therefore favor the reference implementation. The margin was 0.01 for both. A paired case bootstrap was used in R `boot` 1.3-28 with 10,000 replicates and a fixed seed per endpoint.

BCa intervals were used when `boot.ci` returned finite ordered limits with finite jackknife acceleration and no warning; percentile intervals were the declared fallback. All eight all-structure endpoints and both spleen-only endpoints met the BCa conditions. Two-sided 95% CIs used the same method. One-sided p values came from BCa interval inversion with `boot.pval` 0.7.0 (`alternative="less"`, `theta_null=0.01`, `pval_precision=0.001`); values at the first grid point are reported as $p < .001$. The primary analysis required one-sided $p < .05$ on both endpoints; because both had to pass, this rule holds the type I error at the nominal level without adjustment. The three secondary analyses used the same margin, with Holm adjustment across their six endpoint tests; each unadjusted one-sided p was at the first one-sided grid point (.0005, half of `pval_precision`), so every adjusted p was at most .003. The other tolerance, organ, and stratum analyses are reported as differences with two-sided 95% BCa intervals.

DSC used the same masks, references, and eligibility rules as NSD: reference-present structures are eligible, an eligible empty prediction scores 0, and structure values are averaged within each case. NSD at 1 and 2 mm used the same bootstrap (Supplemental Table S8).

**Direct agreement between implementations and per-case tails**

Direct agreement was the DSC between the TS-Portable mask and the reference implementation's mask for the same structure and case, computed on the full three-dimensional masks; pairs in which both masks were empty were excluded, and a pair in which only one was empty scored 0 (Supplemental Table S6). The per-case tail criterion took, for each analysis, the distribution of the per-case paired difference (reference implementation minus TS-Portable) in mean DSC and in mean NSD@3 mm and reports its median, 90th and 95th percentiles, maximum, and the number of cases above 0.005 and 0.01 (Supplemental Table S7).

**Volume-stratified analysis**

Reference volume was voxel count times voxel volume for every reference-present structure instance. Each structure was assigned once, by its median reference volume within a dataset, to one of four strata with cutpoints 25, 75, and 185 mL. No standard size categories exist for these structures; the cutpoints are therefore the rounded quartiles of instance volume over the development set's 15 abdominal structures (25.6, 74.0, and 185.2 mL; the AMOS quartiles were 23.3, 72.5, and 156.2 mL, and the same cuts were used for AMOS). On AMOS the strata were the right and left adrenal glands, esophagus, gallbladder, and prostate (<25 mL); duodenum, inferior vena cava, and pancreas (25–75 mL); bladder, aorta, and right and left kidneys (75–185 mL); and spleen, stomach, and liver (>185 mL). The development set had 56, 29, 10, and 22 structures per stratum. Within each stratum, the paired difference in DSC and NSD@3 mm was pooled over all instances, with a 10,000-resample case bootstrap and BCa intervals. A second grouping compared hollow organs (gastrointestinal tract, bladder, gallbladder) with all other structures. Spearman correlation between the per-structure mean difference and log median volume was computed per dataset and metric. Results are in Supplemental Table S11 and Supplemental Figure S3.

**Runtime benchmarking on the workstation**

The workstation had a 12-core, 24-thread AMD Ryzen Threadripper PRO 7945WX CPU, 128 GB RAM, and an NVIDIA RTX A6000 GPU. The fp32 ONNX arms ran through ONNX Runtime with the OpenVINO execution provider (CPU) and the CUDA execution provider (GPU). Every arm read the same NIfTI files with default thread counts and settings (no test-time augmentation; the reference implementation's sliding-window step, which TS-Portable reproduces). Wall time was measured cold and end to end, from process launch through output-mask writing (input reading, resampling, sliding-window inference and reassembly, postprocessing, mask writing), one case per process, with model loading and compilation timed (no compiled-model cache) and file caches not cleared. Memory was the per-case peak proportional set size over the process tree.

TS-Portable INT8 CPU times were paired by case with each of the four comparator arms on all 100 AMOS scans. A paired case bootstrap with 10,000 resamples (R 4.1.2, `boot` 1.3-28) gave the ratio of the two arms' median cold wall times (the comparator's median over cases divided by TS-Portable's), with BCa 95% CIs for the ratio. Each arm was timed once per case; the intervals therefore reflect case sampling, not run-to-run variation. Ratios are in Table 3 of the main text; software versions of the five arms are in Supplemental Table S3.

**PACS reading station: runtime and spleen volumetry**

The reading station (hardware as in the main text) ran Windows 11 Enterprise with NVIDIA driver 516.25. Each configuration ran one warm-up case and then the 100 cases in sequence, without a compiled-model cache; process-tree resources were sampled every 250 ms. The TS-Portable executable bundled ONNX Runtime with OpenVINO; the user selects a DICOM series or NIfTI file, and the executable writes the mask and a per-structure volume table beside it. No CUDA run fell back to the CPU.

The station ran TotalSegmentator 2.4.0 with PyTorch 2.0.1 and CUDA 11.7 because its driver could not run the CUDA stack that version 2.12.0 requires; the driver of a clinical station was not changed. Its timings describe this deployment, not the reference implementation's GPU performance on supported hardware.

Spleen volume was voxel count times voxel volume on the input grid, for the manual AMOS mask (label 1), the reference implementation's mask, and TS-Portable's mask. The agreement statistics of the main text were also computed for the full 117-label mode on the workstation and for the fp32 ONNX models (Supplemental Table S12).

**Duke Spleen Data Set: spleen volumetry from DICOM**

The Duke Spleen Data Set (Zenodo 7636640) provides 29 axial post-contrast CT series with manual spleen masks stored as binary DICOM images, one per image slice. Each image series and its mask were converted to NIfTI on the image grid (attenuation in Hounsfield units after the rescale slope and intercept; orientation from the image position and orientation attributes). In one series (DSDS_0021) the mask slices were offset from the image slices: 60% of mask voxels fell on soft-tissue attenuation (0–250 HU), against 96%–100% in the other series, and the mask matched the anatomy only after a shift of seven slices. Because realigning it would alter a public reference standard, the series was excluded from all analyses.

The series were distributed with JPEG Lossless (12), JPEG 2000 (7), or uncompressed (9) encoding and were given to TS-Portable unchanged; its DICOM reader and the conversion above yielded identical voxel values and geometry for every series. On the reading station, the DICOM and NIfTI inputs were run as two configurations, each with one warm-up case, under the protocol of the AMOS runs. The reference implementation 2.12.0 ran in spleen-only mode from the NIfTI files on the workstation GPU; only its masks were used. DSC and NSD against the manual masks were computed as in the AMOS analyses and are descriptive (Supplemental Table S13).

**Reference implementation version 2.4.0 versus 2.12.0**

The PACS reading station ran TotalSegmentator 2.4.0 (PyTorch 2.0.1, CUDA 11.7), whereas the workstation comparisons used 2.12.0. To check that the version does not affect the comparison, both versions were run on the workstation's RTX A6000 over the 100 AMOS scans, in spleen-only mode and in full 117-label mode, from the same input files and through the same timing harness, each in its own Python environment (2.4.0 with nnU-Net 2.2.1 and PyTorch 2.0.1; 2.12.0 with nnU-Net 2.6.4 and PyTorch 2.10.0). The weight files that 2.4.0 downloaded for the six models used (five 1.5-mm models and the 6-mm localization model) were byte-identical to those of 2.12.0 (MD5). Results are in Supplemental Table S4; the two versions differ in the code around the model, not in the model, and their outputs differ only at mask boundaries.

**Supplemental Tables**

**Supplemental Table S1. Cohorts and imaging characteristics**

**A. Cohorts**

| Cohort or analysis set | Source | n (CT scans)* | Age, y† | Sex (men/women) | Vendors / sites | Coverage and notes |
|---|---|---|---|---|---|---|
| External AMOS validation set | AMOS validation (Zenodo 7262581) | 100 | 53 ± 16 (13–94) | 53 / 47†† | Siemens (50), Toshiba (27), Philips (23); 2 sites | 15 abdominal organs versus manual reference standard: spleen, right and left kidney, gallbladder, esophagus, liver, stomach, aorta, inferior vena cava, pancreas, right and left adrenal gland, duodenum, urinary bladder, prostate; primary analysis at 1.5 mm and secondary at 3 mm; spleen volumetry |
| TotalSegmentator development set | TotalSegmentator dataset, small release (Zenodo 10047263) | 102 | 67 ± 13 (23–91) | 63 / 38‡ | Siemens (100), GE HealthCare (2) | All 117 labels; 98 of the 102 scans were training data for TotalSegmentator§; quantization fidelity audit at both resolutions |

| Cohort or analysis set | Source | n (CT scans)* | Age, y† | Sex (men/women) | Vendors / sites | Coverage and notes |
|---|---|---|---|---|---|---|
| Duke Spleen Data Set, CT | Duke Spleen Data Set (Zenodo 7636640) | 28 | Not available‖ | Not available‖ | GE (13), Siemens (13), Philips (1), Toshiba (1)#; 1 site | Spleen versus manual reference standard in known or suspected chronic liver disease; spleen volumetry on the reading station from DICOM and NIfTI input; 1 of 29 series excluded (reference mask not aligned with the image series) |
| CT-ORG quantization set | CT-ORG | 28 | Not available‖ | Not available‖ | Multicenter | Quantization development only, with no evaluation: calibration (volumes 0–7), accuracy selection (8–17), and independent patch testing (18–27); disjoint from both evaluation cohorts |

* One CT scan per patient. † Mean ± standard deviation (range). ‡ Sex unspecified for one participant. § Performance in this cohort is optimistic and not comparable with the external AMOS values; the comparison between implementations is unaffected because both run the same weights. †† Assigned from the pelvic anatomy (see AMOS pelvic label and sex assignment); 62 / 38 as recorded. ‖ Not in the public metadata. # From the scanner model names in the image headers. Examination dates were unavailable for all four datasets.

**B. Image geometry, read from the image headers**

| Cohort | In-plane pixel spacing, mm | Slice spacing, mm | Slices per scan | Contrast phase |
|---|---|---|---|---|
| External AMOS validation set | 0.67 (0.45–1.04) | 5.0 (1.25–5.0); 5.0 mm in 62, 2.0–2.5 mm in 36, 1.25 mm in 2 | 118 (74–353) | Not in the public metadata |
| TotalSegmentator development set | 1.5 (all)¶ | 1.5 (all)¶ | 400 (90–549) | Not in the public metadata |
| Duke Spleen Data Set, CT | 0.78 (0.63–0.98) | 5.0 (2.5–5.0); 5.0 mm in 20, 3.0 mm in 6, 2.5 mm in 2 | 86 (34–150) | Post-contrast |
| CT-ORG quantization set | 0.77 (0.65–1.37) | 1.0 (0.7–5.0) | 539 (75–861) | Not in the public metadata |

Values are median (range). ¶The public release of the TotalSegmentator dataset is resampled to 1.5-mm isotropic voxels.

**Supplemental Table S2. Quantization per model: convolutional layers and fidelity achieved**

**A. Convolutional layers quantized to INT8 and kept in fp32**

| Model | Convolutions, INT8 / total | Transposed convolutions, INT8 / total | Kept in fp32 by design* | Restored to fp32 by accuracy control | Share quantized |
|---|---|---|---|---|---|
| 1.5 mm, organs | 18 / 23 | 4 / 5 | 2 | 4 | 22 of 28 (79%) |
| 1.5 mm, vertebrae | 20 / 23 | 4 / 5 | 2 | 2 | 24 of 28 (86%) |
| 1.5 mm, cardiac | 19 / 23 | 3 / 5 | 2 | 4 | 22 of 28 (79%) |
| 1.5 mm, muscles | 21 / 23 | 5 / 5 | 2 | 0 | 26 of 28 (93%) |
| 1.5 mm, ribs | 21 / 23 | 5 / 5 | 2 | 0 | 26 of 28 (93%) |
| 3 mm | 15 / 19 | 2 / 4 | 2 | 4 | 17 of 23 (74%) |
| 6 mm (localization) | 14 / 19 | 3 / 4 | 2 | 4 | 17 of 23 (74%) |
| All seven models | 128 / 153 | 26 / 33 | 14 | 18 | 154 of 186 (83%) |

Counted on the released ONNX graphs: a convolution is INT8 when its weight input comes from a dequantize operation. *The first encoder convolution and the final segmentation convolution of each model were excluded from quantization. Non-convolutional operations (normalization, activation, resampling) stayed in fp32 in every model. fp32 = 32-bit floating-point; INT8 = 8-bit integer.

**B. Fidelity to the fp32 model on CT-ORG patches**

| Model | Layers restored | Selection patches, quantization runtime: initial → final | Target met (≥ 0.995) | Held-out patches, quantization runtime: initial → final | Selection patches, deployed runtime | Held-out patches, deployed runtime |
|---|---|---|---|---|---|---|
| 1.5 mm, organs | 4 | 0.9826 → 0.9889 | No | 0.9713 → 0.9678 | 0.9880 | 0.9682 |
| 1.5 mm, vertebrae | 2 | 0.9931 → 0.9952 | Yes | 0.9734 → 0.9748 | 0.9941 | 0.9739 |
| 1.5 mm, cardiac | 4 | 0.9881 → 0.9957 | Yes | 0.9800 → 0.9865 | 0.9957 | 0.9867 |
| 1.5 mm, muscles | 0 | 0.9969 → 0.9969 | Yes | 0.9692 → 0.9692 | 0.9969 | 0.9702 |
| 1.5 mm, ribs | 0 | 0.9964 → 0.9964 | Yes | 0.9580 → 0.9580 | 0.9865 | 0.9647 |
| 3 mm | 4 | 0.9833 → 0.9909 | No | 0.9473 → 0.9574 | 0.9910 | 0.9484 |
| 6 mm (localization) | 4 | 0.9799 → 0.9840 | No | 0.9706 → 0.9774 | 0.9843 | 0.9778 |

Fidelity is the mean over patches of the mean Dice, across labels, between the quantized and fp32 predictions; a label present in only one of the two predictions scores 0, and the fp32 model scores 1. The 0.005 limit on fidelity loss therefore corresponds to a fidelity of at least 0.995. Initial = the model with all eligible operations quantized, before any restoration (quantization without accuracy control); final = the released model. The quantization runtime was mainline ONNX Runtime 1.27.0 (CPU), on which the accuracy-control loop ran; the deployed runtime was `onnxruntime-openvino` 1.24.1 with the released UINT8-encoded files. Twenty selection patches (volumes 8–17) and twenty held-out patches (volumes 18–27) per model, two per volume, chosen as the most foreground-rich candidate patches. The 6-mm, 3-mm, and 1.5-mm organ models stopped at the four-iteration limit without reaching the target and were kept as they stood. fp32 = 32-bit floating-point.

**Supplemental Table S3. Software stages and versions**

| Stage | Component | Version | Role |
|---|---|---|---|
| Model export | TotalSegmentator weights | 2.12.0 task `total` (nnU-Net 2.6.4) | source fp32 weights, unchanged |
| Model export | ONNX | 1.22.0 | fp32 graph format |
| Quantization | NNCF `quantize_with_accuracy_control` | 3.2.0 | accuracy-controlled INT8 candidate generation |
| Quantization | ONNX Runtime (mainline) | 1.27.0 | reference inference inside the NNCF loop (signed-INT8 zero points) |
| Deployment | `onnxruntime-openvino` / OpenVINO | 1.24.1 / 2025.4.1 | executes the released UINT8 QDQ graphs; all TS-Portable CPU results |
| Deployment | ONNX Runtime CUDA execution provider | 1.22.0, cuDNN 9 | GPU arm of the runtime comparison, byte-identical fp32 models |
| Deployment | TS-Portable executable (Rust) | 0.1.0 | reads input, runs inference, writes masks and volume table; distributed as one folder holding the executable, the ONNX Runtime and OpenVINO libraries, and the seven INT8 model files (about 350 MB, of which the models are 212 MB), run without installation |
| Reference implementation | TotalSegmentator / PyTorch | 2.12.0 / 2.10.0 (workstation); 2.4.0 / 2.0.1, CUDA 11.7 (PACS station) | comparator arms |
| Analysis | surface-distance (Google DeepMind) | 0.1 | NSD (surface Dice) by `compute_surface_dice_at_tolerance` on surfels in physical voxel spacing |
| Analysis | R / `boot` / `boot.pval` | 4.1.2 / 1.3-28 / 0.7.0 | paired case bootstrap, BCa intervals, interval-inversion p values |

**Supplemental Table S4. Reference implementation 2.4.0 versus 2.12.0 on the workstation GPU, 100 AMOS cases**

| Mode | Quantity | 2.4.0 | 2.12.0 | Agreement between versions |
|---|---|---|---|---|
| Spleen-only | Spleen volume vs manual reference, CCC | 0.989 | 0.989 | CCC 0.99999; relative bias 0.00%; LOA −0.35% to 0.34%; no case differing by more than 2% |
| Spleen-only | Spleen volume vs manual reference, relative bias (LOA) | −6.9% (−32.9 to 19.2) | −6.8% (−33.0 to 19.3) | |
| Spleen-only | Spleen DSC vs manual reference, mean | 0.9431 | 0.9433 | Direct DSC between versions: median 0.9999, mean 0.9987, minimum 0.983; 10 of 100 masks identical |
| Spleen-only | Wall time per case, median (90th percentile), s | 14.8 (16.8) | 16.4 (18.7) | Same session, cases in the same order |
| Full 117-label | Per-case mean DSC over the 15 organs vs manual reference | 0.8496 | 0.8495 | Difference 2.12.0 minus 2.4.0: mean −0.00007, largest absolute 0.0070 |

| Mode | Quantity | 2.4.0 | 2.12.0 | Agreement between versions |
|---|---|---|---|---|
| Full 117-label | Direct DSC between versions, per organ, mean (minimum) | — | — | Spleen 0.9996 (0.994); liver 0.9994 (0.993); kidneys 0.9995–0.9998 (0.995); lowest means right adrenal gland 0.9955 (0.934), esophagus 0.9961 (0.959), and gallbladder 0.9965 (0.883) |
| Full 117-label | Wall time per case, median (90th percentile), s | 51.7 (73.7) | 61.5 (Table 3) | Different sessions; the 2.12.0 value is the Table 3 run |

Spleen-only mode uses the 6-mm localization and 1.5-mm organ models. Volumes are voxel count times voxel volume; relative differences are 2.4.0 minus 2.12.0 as a percentage of the pair mean. The 2.4.0 masks produced on the reading station's CPU and on the workstation's GPU agreed with CCC 1.00000 and limits of −0.3% to 0.2%, so neither hardware nor operating system moved the reference implementation's volumes. CCC = Lin's concordance correlation coefficient; DSC = Dice similarity coefficient; LOA = 95% limits of agreement.

**Supplemental Table S5. ONNX conversion fidelity**

| Cohort and resolution | n | fp32 ONNX | Reference implementation | Difference, NSD@3 mm* | Difference, DSC* |
|---|---|---|---|---|---|
| External AMOS, 1.5 mm | 100 | 0.9088 | 0.9088 | −0.00001 | −0.00002 |
| External AMOS, 3 mm | 100 | 0.8857 | 0.8856 | −0.00006 | −0.00012 |
| Development set, 1.5 mm | 102 | 0.9550 | 0.9550 | −0.00002 | −0.00001 |
| Development set, 3 mm | 102 | 0.9571 | 0.9571 | 0.00001 | −0.00006 |

Per-case mean NSD@3 mm of the unquantized fp32 ONNX models in TS-Portable and of the reference implementation, with the differences in NSD@3 mm and DSC. *Reference implementation minus fp32 ONNX, the same direction as the main analyses; positive values favor the reference implementation.

**Supplemental Table S6. Direct mask-to-mask agreement between TS-Portable and the reference implementation on external AMOS**

| Organ | 1.5 mm: n | 1.5 mm: mean DSC | 1.5 mm: lowest case | 3 mm: n | 3 mm: mean DSC | 3 mm: lowest case |
|---|---|---|---|---|---|---|
| Liver | 100 | 0.9986 | 0.9953 | 100 | 0.9960 | 0.9837 |
| Stomach | 99 | 0.9970 | 0.9733 | 99 | 0.9914 | 0.9085 |
| Spleen | 100 | 0.9983 | 0.9869 | 100 | 0.9952 | 0.9675 |
| Left kidney | 100 | 0.9987 | 0.9915 | 100 | 0.9959 | 0.9587 |
| Right kidney | 98 | 0.9986 | 0.9936 | 98 | 0.9956 | 0.9676 |
| Aorta | 100 | 0.9982 | 0.9935 | 100 | 0.9945 | 0.9814 |
| Urinary bladder | 99 | 0.9942 | 0.9506 | 99 | 0.9842 | 0.8827 |
| Pancreas | 100 | 0.9953 | 0.9773 | 100 | 0.9871 | 0.9554 |
| Inferior vena cava | 100 | 0.9915 | 0.9769 | 100 | 0.9861 | 0.9605 |
| Duodenum | 99 | 0.9930 | 0.9603 | 99 | 0.9766 | 0.9135 |
| Prostate | 52 | 0.9944 | 0.9815 | 52 | 0.9857 | 0.9533 |
| Gallbladder | 92 | 0.9941 | 0.9556 | 87 | 0.9824 | 0.8258 |
| Esophagus | 100 | 0.9961 | 0.9825 | 100 | 0.9859 | 0.9548 |
| Left adrenal gland | 100 | 0.9937 | 0.9524 | 99 | 0.9784 | 0.8189 |
| Right adrenal gland | 100 | 0.9923 | 0.9231 | 100 | 0.9781 | 0.8900 |

DSC between the two implementations' masks for the same organ and case, over the n organ–case reference-present pairs in which at least one mask was nonempty; the mean over organs was 0.9956 at 1.5 mm and 0.9875 at 3 mm. Across all pairs, the 5th percentile was 0.9868 (1.5 mm) and 0.9649 (3 mm), and the lowest pair was 0.9231 (a right adrenal gland) and 0.8189 (a left adrenal gland). In the development set, the mean over the 117 labels was 0.9957 at 1.5 mm (lowest label mean, skull, 0.9681) and 0.9920 at 3 mm (lowest, C6 vertebra, 0.9675), with 5th percentiles of 0.9893 and 0.9813; the lowest pairs with two nonempty masks were a left 12th rib (0.5371) and a right middle lung lobe (0.7692), and 2 (1.5 mm) and 1 (3 mm) pairs were empty for TS-Portable only. DSC = Dice similarity coefficient.

**Supplemental Table S7. Distribution of the paired difference per case (tail criterion) and per organ and case**

**A. Per case (difference in the per-case mean)**

| Analysis | Metric | Median | 90th percentile | 95th percentile | Maximum | Cases > 0.005 | Cases > 0.01 |
|---|---|---|---|---|---|---|---|
| External AMOS, 1.5 mm | DSC | 0.0002 | 0.0009 | 0.0012 | 0.0023 | 0 | 0 |
| External AMOS, 1.5 mm | NSD@3 mm | 0.0001 | 0.0011 | 0.0016 | 0.0025 | 0 | 0 |
| External AMOS, 3 mm | DSC | 0.0001 | 0.0023 | 0.0038 | 0.0092 | 3 | 0 |
| External AMOS, 3 mm | NSD@3 mm | −0.0001 | 0.0027 | 0.0037 | 0.0141 | 4 | 1 |
| Development set, 1.5 mm | DSC | 0.0001 | 0.0005 | 0.0007 | 0.0020 | 0 | 0 |
| Development set, 1.5 mm | NSD@3 mm | 0.0000 | 0.0002 | 0.0005 | 0.0019 | 0 | 0 |
| Development set, 3 mm | DSC | 0.0003 | 0.0008 | 0.0013 | 0.0072 | 1 | 0 |
| Development set, 3 mm | NSD@3 mm | 0.0001 | 0.0006 | 0.0008 | 0.0129 | 1 | 1 |

Per-case paired difference in the per-case mean metric, reference implementation minus TS-Portable; positive values favor the reference implementation. The two cases above 0.01 at 3 mm are one AMOS case (gallbladder and adrenal differences) and one development-set case. DSC = Dice similarity coefficient; NSD@3 mm = normalized surface distance at a 3-mm tolerance.

**B. Per organ and case, against the manual reference**

| Analysis | Metric | Pairs | Median | 95th percentile | 99th percentile | Largest loss | Largest gain | Losses > 0.01 | Losses > 0.05 | Losses > 0.10 | Gains > 0.01 |
|---|---|---|---|---|---|---|---|---|---|---|---|
| External AMOS, 1.5 mm | DSC | 1,440 | 0.0001 | 0.0036 | 0.0074 | 0.028 | 0.038 | 6 | 0 | 0 | 11 |
| External AMOS, 1.5 mm | NSD@3 mm | 1,440 | 0.0000 | 0.0044 | 0.0101 | 0.041 | 0.030 | 15 | 0 | 0 | 17 |
| External AMOS, 3 mm | DSC | 1,440 | 0.0000 | 0.0082 | 0.0204 | 0.132 | 0.065 | 50 | 4 | 1 | 60 |
| External AMOS, 3 mm | NSD@3 mm | 1,440 | 0.0000 | 0.0113 | 0.0232 | 0.170 | 0.121 | 86 | 4 | 1 | 77 |
| Development set, 1.5 mm | DSC | 9,299 | 0.0001 | 0.0023 | 0.0065 | 0.114 | 0.057 | 49 | 3 | 2 | 45 |
| Development set, 1.5 mm | NSD@3 mm | 9,299 | 0.0000 | 0.0015 | 0.0054 | 0.157 | 0.059 | 47 | 3 | 1 | 44 |
| Development set, 3 mm | DSC | 9,299 | 0.0001 | 0.0040 | 0.0096 | 0.444 | 0.061 | 86 | 3 | 2 | 47 |
| Development set, 3 mm | NSD@3 mm | 9,299 | 0.0000 | 0.0032 | 0.0095 | 0.823 | 0.060 | 84 | 2 | 1 | 61 |

Paired difference for each reference-present organ–case pair (structure–case pair in the development set), reference implementation minus TS-Portable, against the manual reference; a loss is a positive difference and a gain a negative one.

Largest losses: AMOS 1.5 mm, a gallbladder (DSC) and a right adrenal gland (NSD@3 mm); AMOS 3 mm, a gallbladder (DSC 0.13, NSD@3 mm 0.17); development set 1.5 mm, the left 12th rib of Supplemental Figure S4; development set 3 mm, a C6 vertebra left empty by TS-Portable only (reference implementation DSC 0.44, NSD@3 mm 0.82). The other two development-set pairs empty for TS-Portable only (a skull and a T8 vertebra, 1.5 mm) scored 0 DSC with both implementations.

**Supplemental Table S8. Surface agreement at 1- and 2-mm NSD tolerances**

| Tolerance | Comparison | n | Reference implementation | TS-Portable | Difference (95% CI)* |
|---|---|---|---|---|---|
| 1 mm | External AMOS, 1.5 mm | 100 | 0.7587 | 0.7581 | 0.00058 (0.00040–0.00077) |
| 1 mm | External AMOS, 3 mm | 100 | 0.6686 | 0.6682 | 0.00030 (−0.00031 to 0.00089) |
| 1 mm | Development set, 1.5 mm | 102 | 0.8410 | 0.8408 | 0.00025 (0.00013–0.00037) |
| 1 mm | Development set, 3 mm | 102 | 0.6004 | 0.6000 | 0.00045 (0.00027–0.00074) |
| 2 mm | External AMOS, 1.5 mm | 100 | 0.8692 | 0.8689 | 0.00030 (0.00014–0.00048) |
| 2 mm | External AMOS, 3 mm | 100 | 0.8328 | 0.8326 | 0.00018 (−0.00047 to 0.00079) |
| 2 mm | Development set, 1.5 mm | 102 | 0.9368 | 0.9368 | 0.00001 (−0.00008 to 0.00010) |
| 2 mm | Development set, 3 mm | 102 | 0.9183 | 0.9178 | 0.00046 (0.00029–0.00087) |

Per-case mean NSD at the stated tolerance. *Reference implementation minus TS-Portable; two-sided 95% BCa interval from 10,000 paired case resamples; none of the eight two-sided upper limits exceeded 0.0009. BCa = bias-corrected and accelerated; CI = confidence interval; NSD = normalized surface distance.

**Supplemental Table S9. Organ-specific DSC and NSD@3 mm on external AMOS**

**A. 1.5-mm model**

| Organ | n | DSC | NSD@3 mm |
|---|---|---|---|
| Liver | 100 | 0.9643/0.9645 | 0.9489/0.9494 |
| Stomach | 99 | 0.9087/0.9085 | 0.8943/0.8938 |
| Spleen | 100 | 0.9497/0.9498 | 0.9651/0.9652 |
| Left kidney | 100 | 0.9472/0.9470 | 0.9778/0.9775 |
| Right kidney | 98 | 0.9466/0.9467 | 0.9791/0.9790 |
| Aorta | 100 | 0.9217/0.9217 | 0.9634/0.9632 |
| Urinary bladder | 99 | 0.8047/0.8042 | 0.7854/0.7854 |
| Pancreas | 100 | 0.8304/0.8304 | 0.8893/0.8889 |
| Inferior vena cava | 100 | 0.8473/0.8505 | 0.9174/0.9200 |
| Duodenum | 99 | 0.7216/0.7214 | 0.7956/0.7954 |
| Prostate | 52 | 0.7763/0.7775 | 0.7723/0.7753 |
| Gallbladder | 93 | 0.8196/0.8190 | 0.8752/0.8745 |
| Esophagus | 100 | 0.8166/0.8172 | 0.9402/0.9404 |
| Left adrenal gland | 100 | 0.7379/0.7384 | 0.9309/0.9311 |
| Right adrenal gland | 100 | 0.7159/0.7147 | 0.9264/0.9261 |

**B. 3-mm model**

| Organ | n | DSC | NSD@3 mm |
|---|---|---|---|
| Liver | 100 | 0.9560/0.9565 | 0.9441/0.9454 |
| Stomach | 99 | 0.8866/0.8855 | 0.8741/0.8730 |
| Spleen | 100 | 0.9374/0.9368 | 0.9671/0.9662 |
| Left kidney | 100 | 0.9280/0.9281 | 0.9746/0.9743 |
| Right kidney | 98 | 0.9261/0.9266 | 0.9715/0.9716 |
| Aorta | 100 | 0.8887/0.8886 | 0.9594/0.9596 |
| Urinary bladder | 99 | 0.7836/0.7849 | 0.7563/0.7599 |
| Pancreas | 100 | 0.7912/0.7907 | 0.8579/0.8567 |
| Inferior vena cava | 100 | 0.8095/0.8103 | 0.9035/0.9035 |
| Duodenum | 99 | 0.6665/0.6678 | 0.7414/0.7422 |
| Prostate | 52 | 0.7636/0.7624 | 0.7639/0.7624 |
| Gallbladder | 93 | 0.7523/0.7521 | 0.8123/0.8118 |
| Esophagus | 100 | 0.7592/0.7591 | 0.9198/0.9193 |
| Left adrenal gland | 100 | 0.6470/0.6484 | 0.8856/0.8863 |
| Right adrenal gland | 100 | 0.6284/0.6273 | 0.8910/0.8890 |

Values are TS-Portable/reference implementation, means over the n eligible cases against the AMOS manual reference; empty predictions score 0. n is below 100 where the organ is absent from the manual reference (right kidney 2, gallbladder 7, stomach, duodenum, and urinary bladder 1 each); the prostate was scored in the 52 men with a labeled prostate (sex assigned from the pelvic anatomy; see AMOS pelvic label and sex assignment). Both implementations left the same cases empty: at 1.5 mm, a gallbladder; at 3 mm, gallbladder 6 and left adrenal gland 1. Paired differences are in Supplemental Table S10. DSC = Dice similarity coefficient; NSD = normalized surface distance.

**Supplemental Table S10. Organ-specific paired differences on external AMOS**

| Model | Organ | n | DSC difference (95% CI)* | NSD@3 mm difference (95% CI)* |
|---|---|---|---|---|
| 1.5 mm | Liver | 100 | 0.00020 (0.00013–0.00026) | 0.00049 (0.00027–0.00079) |
| 1.5 mm | Stomach | 99 | −0.00026 (−0.00085 to 0.00003) | −0.00050 (−0.00117 to −0.00006) |
| 1.5 mm | Spleen | 100 | 0.00005 (−0.00007 to 0.00022) | 0.00004 (−0.00032 to 0.00040) |
| 1.5 mm | Left kidney | 100 | −0.00019 (−0.00044 to −0.00005) | −0.00029 (−0.00073 to −0.00006) |
| 1.5 mm | Right kidney | 98 | 0.00007 (−0.00005 to 0.00018) | −0.00009 (−0.00031 to 0.00012) |
| 1.5 mm | Aorta | 100 | −0.00006 (−0.00018 to 0.00007) | −0.00018 (−0.00051 to 0.00011) |
| 1.5 mm | Urinary bladder | 99 | −0.00042 (−0.00188 to 0.00030) | −0.00005 (−0.00115 to 0.00072) |
| 1.5 mm | Pancreas | 100 | −0.00003 (−0.00041 to 0.00023) | −0.00047 (−0.00087 to −0.00015) |
| 1.5 mm | Inferior vena cava | 100 | 0.00317 (0.00268–0.00382) | 0.00252 (0.00196–0.00317) |
| 1.5 mm | Duodenum | 99 | −0.00017 (−0.00067 to 0.00024) | −0.00027 (−0.00079 to 0.00023) |
| 1.5 mm | Prostate | 52 | 0.00126 (0.00068–0.00198) | 0.00298 (0.00161–0.00455) |
| 1.5 mm | Gallbladder | 93 | −0.00058 (−0.00179 to 0.00047) | −0.00077 (−0.00189 to 0.00017) |
| 1.5 mm | Esophagus | 100 | 0.00068 (0.00019–0.00102) | 0.00014 (−0.00051 to 0.00047) |
| 1.5 mm | Left adrenal gland | 100 | 0.00049 (−0.00014 to 0.00128) | 0.00018 (−0.00082 to 0.00128) |
| 1.5 mm | Right adrenal gland | 100 | −0.00126 (−0.00200 to −0.00060) | −0.00023 (−0.00092 to 0.00126) |
| 3 mm | Liver | 100 | 0.00054 (0.00037–0.00087) | 0.00131 (0.00070–0.00224) |
| 3 mm | Stomach | 99 | −0.00110 (−0.00257 to −0.00037) | −0.00111 (−0.00237 to 0.00002) |
| 3 mm | Spleen | 100 | −0.00059 (−0.00159 to −0.00013) | −0.00082 (−0.00232 to 0.00028) |
| 3 mm | Left kidney | 100 | 0.00007 (−0.00032 to 0.00125) | −0.00038 (−0.00144 to 0.00011) |
| 3 mm | Right kidney | 98 | 0.00048 (−0.00005 to 0.00142) | 0.00008 (−0.00164 to 0.00080) |
| 3 mm | Aorta | 100 | −0.00014 (−0.00049 to 0.00009) | 0.00015 (−0.00019 to 0.00040) |

| Model | Organ | n | DSC difference (95% CI)* | NSD@3 mm difference (95% CI)* |
|---|---|---|---|---|
| 3 mm | Urinary bladder | 99 | 0.00133 (−0.00119 to 0.00380) | 0.00362 (0.00087–0.00592) |
| 3 mm | Pancreas | 100 | −0.00042 (−0.00135 to 0.00058) | −0.00118 (−0.00222 to −0.00020) |
| 3 mm | Inferior vena cava | 100 | 0.00082 (−0.00012 to 0.00168) | 0.00005 (−0.00110 to 0.00117) |
| 3 mm | Duodenum | 99 | 0.00131 (−0.00008 to 0.00300) | 0.00080 (−0.00093 to 0.00242) |
| 3 mm | Prostate | 52 | −0.00125 (−0.00290 to 0.00064) | −0.00143 (−0.00416 to 0.00183) |
| 3 mm | Gallbladder | 93 | −0.00021 (−0.00232 to 0.00501) | −0.00055 (−0.00364 to 0.00583) |
| 3 mm | Esophagus | 100 | −0.00004 (−0.00092 to 0.00074) | −0.00054 (−0.00139 to 0.00026) |
| 3 mm | Left adrenal gland | 100 | 0.00136 (−0.00060 to 0.00444) | 0.00078 (−0.00102 to 0.00324) |
| 3 mm | Right adrenal gland | 100 | −0.00111 (−0.00391 to 0.00128) | −0.00201 (−0.00660 to 0.00081) |

*Reference implementation minus TS-Portable, mean over the n eligible cases with two-sided 95% BCa interval from 10,000 paired case resamples; positive values favor the reference implementation. BCa = bias-corrected and accelerated; CI = confidence interval; DSC = Dice similarity coefficient; NSD = normalized surface distance.

**Supplemental Table S11. Quantization effect by structure volume stratum (1.5-mm models)**

**A. Development set**

| Stratum | Structures | Instances | DSC difference (95% CI)* | NSD@3 mm difference (95% CI)* |
|---|---|---|---|---|
| <25 mL | 56 | 4,333 | 0.00001 (−0.00012 to 0.00017) | 0.00002 (−0.00009 to 0.00018) |
| 25–75 mL | 29 | 2,247 | 0.00016 (0.00010–0.00025) | 0.00002 (−0.00004 to 0.00010) |
| 75–185 mL | 10 | 827 | 0.00010 (−0.00005 to 0.00022) | −0.00001 (−0.00014 to 0.00011) |
| >185 mL | 22 | 1,892 | 0.00021 (0.00010–0.00034) | 0.00008 (0.00001–0.00016) |
| Hollow organs | 7 | 611 | 0.00007 (−0.00030 to 0.00023) | 0.00018 (0.00003–0.00036) |
| All other structures | 110 | 8,688 | 0.00010 (0.00002–0.00019) | 0.00002 (−0.00004 to 0.00011) |

**B. External AMOS**

| Stratum | Structures | Instances | DSC difference (95% CI)* | NSD@3 mm difference (95% CI)* |
|---|---|---|---|---|
| <25 mL | 5 | 445 | 0.00001 (−0.00034 to 0.00034) | 0.00021 (−0.00021 to 0.00065) |
| 25–75 mL | 3 | 299 | 0.00099 (0.00069–0.00129) | 0.00060 (0.00028–0.00089) |
| 75–185 mL | 4 | 397 | −0.00015 (−0.00050 to 0.00005) | −0.00015 (−0.00045 to 0.00008) |
| >185 mL | 3 | 299 | 0.00000 (−0.00019 to 0.00011) | 0.00001 (−0.00024 to 0.00021) |
| Hollow organs | 5 | 490 | −0.00014 (−0.00048 to 0.00014) | −0.00028 (−0.00067 to 0.00001) |
| All other structures | 10 | 950 | 0.00033 (0.00020–0.00045) | 0.00037 (0.00019–0.00056) |

Structures were assigned to strata by median reference volume. Differences are reference implementation minus TS-Portable, pooled over all instances of the stratum's structures, with two-sided 95% BCa intervals from a 10,000-resample case bootstrap. Spearman correlation of the per-structure difference with log median volume: DSC 0.11 (development set) and −0.03 (AMOS); NSD@3 mm 0.05 and −0.09. Hollow organs: gastrointestinal tract, urinary bladder, gallbladder.
*Positive values favor the reference implementation.

**Supplemental Table S12. Spleen volume agreement, all comparisons, 100 external AMOS cases**

| Setting | Comparison | CCC (95% BCa CI) | Relative bias, % | 95% LOA, % | Absolute bias, mL |
|---|---|---|---|---|---|
| PACS station, spleen-only mode | TS-Portable vs manual | 0.989 (0.978–0.994) | −6.8 | −28.5 to 15.0 | −13.8 |
| PACS station, spleen-only mode | Reference implementation 2.4.0 (CPU) vs manual | 0.989 (0.978–0.994) | −6.8 | −32.8 to 19.1 | −13.5 |
| PACS station, spleen-only mode | TS-Portable vs reference implementation 2.4.0 (CPU) | 0.9999 (0.9998–1.0000) | 0.2 | −7.2 to 7.6 | −0.2 |
| PACS station, spleen-only mode | Reference implementation CPU vs CUDA | 1.0000 (1.0000–1.0000) | 0.0 | −0.2 to 0.3 | 0.0 |
| Workstation, full 117-label mode | TS-Portable vs manual | 0.988 (0.975–0.993) | −5.9 | −16.7 to 5.0 | −14.1 |
| Workstation, full 117-label mode | Reference implementation 2.12.0 vs manual | 0.988 (0.976–0.993) | −5.8 | −16.5 to 4.9 | −13.9 |
| Workstation, full 117-label mode | TS-Portable vs reference implementation 2.12.0 | 1.0000 (1.0000–1.0000) | −0.1 | −0.5 to 0.3 | −0.1 |
| Workstation, full 117-label mode | fp32 ONNX vs reference implementation 2.12.0 | 1.0000 (1.0000–1.0000) | 0.0 | 0.0 to 0.0 | 0.0 |

Manual spleen volumes: median 187 mL (interquartile range, 139–261 mL; range, 16–954 mL). Relative bias and limits of agreement (mean ± 1.96 SD of the relative differences) are the first-named minus the second-named volume as a percentage of the pair mean; CCC intervals are two-sided 95% BCa intervals from 10,000 case resamples; bootstrap intervals for the bias and limits of the spleen-only-mode rows are in Table 5. Because one case dominates the standard deviation, the empirical distribution is also given: the 2.5th and 97.5th percentiles of the relative differences were −20.6% and 0.9% for TS-Portable vs manual and −1.4% and 1.5% for TS-Portable vs reference implementation; the median absolute difference was 8.9 mL (5.0%) and 0.3 mL (0.2%); and 13, 3, and 1 TS-Portable volumes differed from the manual volume by more than 10%, 20%, and 50% (all percentages of the pair mean). The smallest spleen (16 mL by manual reference; 4.9 mL by TS-Portable and 3.4 mL by the reference implementation in spleen-only mode) determines most of the width of these limits: without that case, the limits are −15.1% to 3.6% against the manual reference and −2.5% to 2.2% between implementations; in full mode the case is measured at 15 mL by both implementations and the limits are unchanged without it. In spleen-only mode the two implementations differed from their full-mode volumes by more than 5% in the same 8 of 100 cases, in the same direction, which is consistent with the wider limits of that mode arising when the 1.5-mm model segments the spleen-only crop, a step TS-Portable reproduces from the reference implementation. In the TS-Portable versus reference implementation row, the relative bias is positive while the absolute bias is slightly negative because the smallest spleen dominates the relative differences. BCa = bias-corrected and accelerated; CCC = Lin's concordance correlation coefficient; CI = confidence interval; LOA = limits of agreement.

**Supplemental Table S13. Duke Spleen Data Set: spleen overlap and runtime on the reading station, 28 CT series**

**A. Overlap of spleen-only-mode masks**

| Measure | TS-Portable vs manual | Reference implementation 2.12.0 vs manual | TS-Portable vs reference implementation 2.12.0 |
|---|---|---|---|
| DSC | 0.961 (0.950–0.967) | 0.962 (0.950–0.967) | 0.998 (0.995–0.999) |
| NSD@1 mm | 0.852 (0.793–0.879) | 0.857 (0.811–0.881) | 0.996 (0.991–0.999) |

| Measure | TS-Portable vs manual | Reference implementation 2.12.0 vs manual | TS-Portable vs reference implementation 2.12.0 |
|---|---|---|---|
| NSD@2 mm | 0.943 (0.915–0.962) | 0.944 (0.924–0.962) | 1.000 (0.999–1.000) |
| NSD@3 mm | 0.981 (0.956–0.991) | 0.981 (0.963–0.991) | 1.000 (1.000–1.000) |

**B. TS-Portable runtime and memory on the reading station by input format**

| Input | Wall time, s | Peak working set, GiB |
|---|---|---|
| NIfTI | 6.4 (4.7–8.4) | 1.58 (1.35–1.96) |
| DICOM | 6.5 (4.9–8.6) | 1.54 (1.36–1.96) |
| Paired difference, DICOM minus NIfTI | 0.12 (0.04–0.25); maximum, 0.54 | — |

Values are median (interquartile range). TS-Portable masks are from DICOM input; masks from NIfTI input were identical in all 28 series. Wall time is from process launch through writing of the output mask, one case per process, after one warm-up case per configuration. DSC = Dice similarity coefficient; NSD = normalized surface distance.

**Supplemental Figures**

**Supplemental Figure S1.** Accuracy-controlled INT8 quantization. Each of the seven fp32 ONNX models was first quantized as a whole, with activation statistics from CT-ORG volumes 0–7, and its fidelity to the fp32 model was measured on a separate accuracy-selection set (volumes 8–17). While the fidelity loss exceeded the chosen threshold of 0.005, the operations most sensitive to quantization were ranked, restored to fp32, and the model reevaluated, for at most four iterations; a model still above the threshold after the last iteration was kept as it stood (achieved values in Supplemental Table S2). Volumes 18–27 were reserved for held-out patch testing. fp32 = 32-bit floating-point; INT8 = 8-bit integer; ONNX = Open Neural Network Exchange; QDQ = quantize–dequantize.

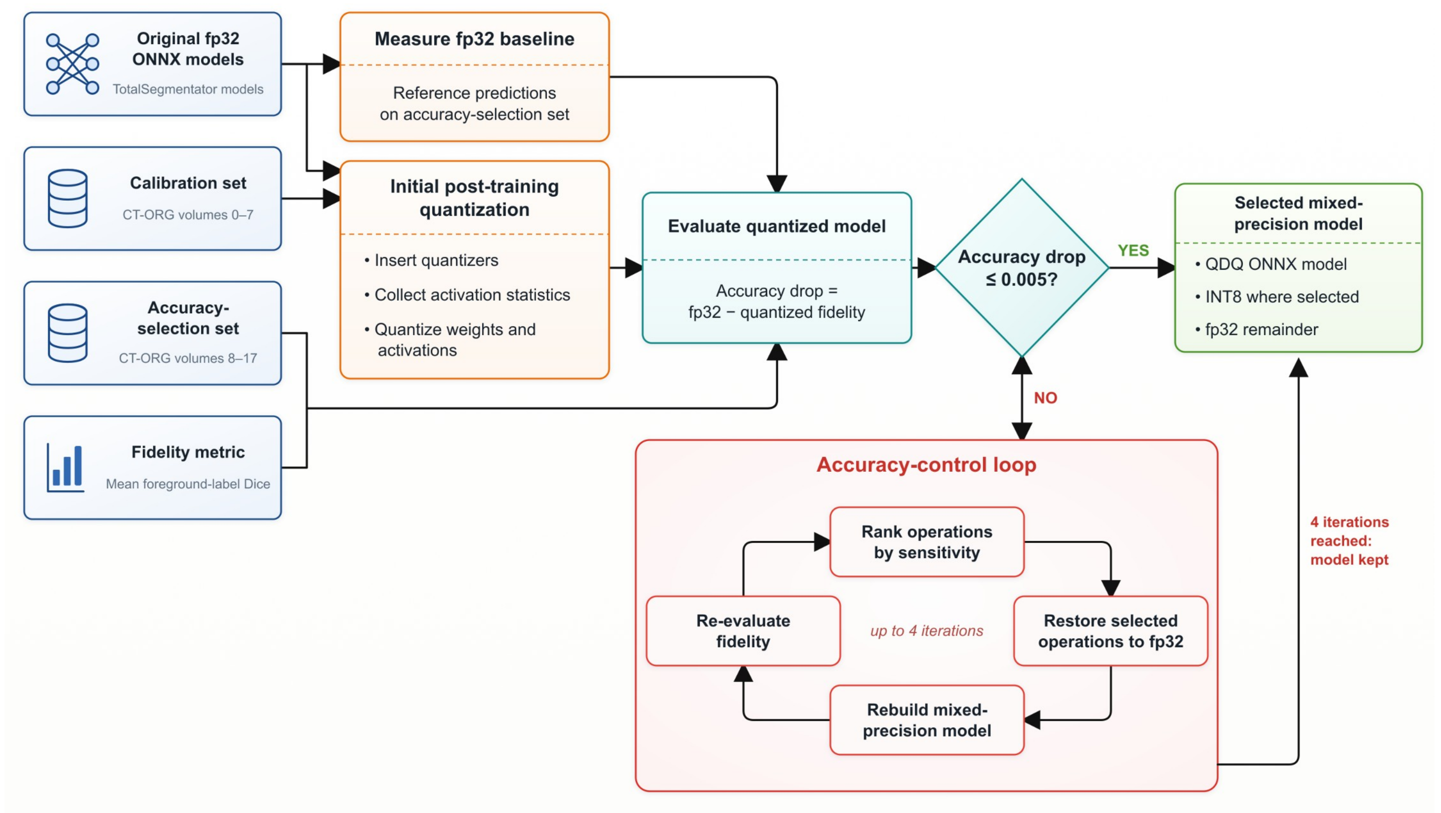

**Supplemental Figure S2.** Further examples from the external AMOS cohort comparing the reference implementation with TS-Portable for the spleen, stomach, liver, and gallbladder, in full 117-label mode. (A) Results at 1.5 mm. (B) Results at 3 mm. Patients, by row: (A) man in his 60s, woman in her 30s, man in his 60s, and man in his 30s; (B) woman in her 30s, woman in her 30s, man in his 30s, and woman in her 30s. The spleen cases are those closest to the cohort's median spleen NSD@3 mm; the stomach, liver, and gallbladder cases were chosen to show differences in either direction and are not a representative sample. NSD@3 mm values, computed on the whole three-dimensional masks, are shown below the segmentations. The 1.5-mm spleen case is the patient shown in Figure 3 of the main text. AMOS = Abdominal Multi-Organ Segmentation; NSD@3 mm = normalized surface distance at a 3-mm tolerance.

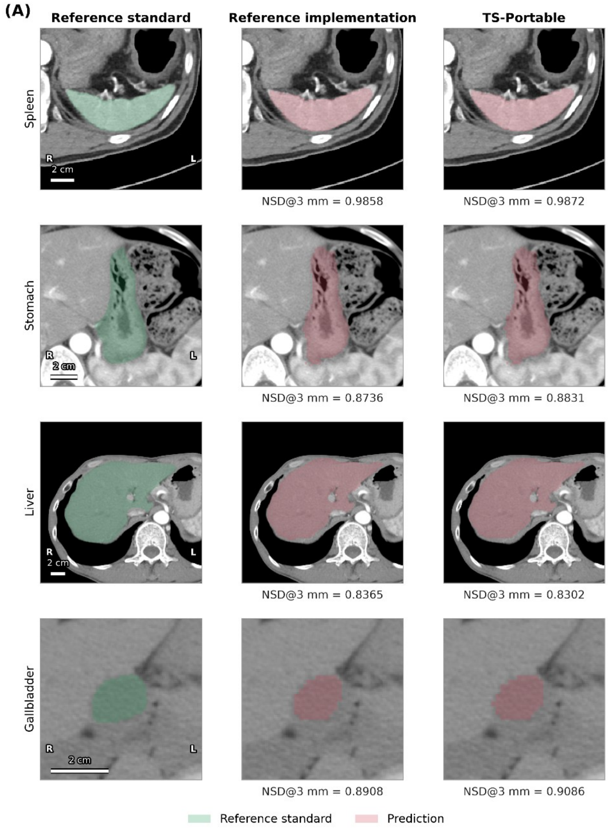

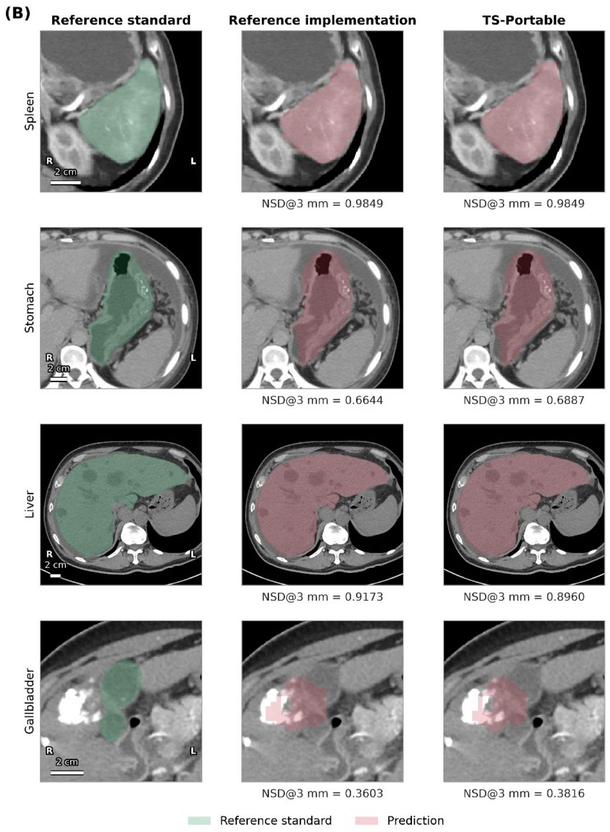
(B)
Reference standard
Reference implementation
TS-Portable
Spleen
R
2 cm
L
NSD@3 mm = 0.9849
NSD@3 mm = 0.9849
Stomach
R
2 cm
L
NSD@3 mm = 0.6644
NSD@3 mm = 0.6887
Liver
R
2 cm
L
NSD@3 mm = 0.9173
NSD@3 mm = 0.8960
Gallbladder
R
2 cm
L
NSD@3 mm = 0.3603
NSD@3 mm = 0.3816
Reference standard
Prediction

**Supplemental Figure S3.** Quantization effect by structure size, 1.5-mm models. Each point is one structure, placed at its median reference volume (logarithmic axis), showing the pooled paired difference (reference implementation minus TS-Portable) across all instances of that structure with its two-sided 95% BCa interval, for DSC (A, B) and NSD@3 mm (C, D) in the 117-label development set (A, C; 102 scans) and the 15 AMOS organs (B, D; 100 scans). The dashed line marks the 0.01 margin, the solid line zero, and the vertical dotted lines the volume strata at 25, 75, and 185 mL. The single structure whose upper bound exceeds the margin, the left 12th rib on NSD@3 mm, is shown in red (Supplemental Figure S4). Stratum-level pooled differences are given in Supplemental Table S11.

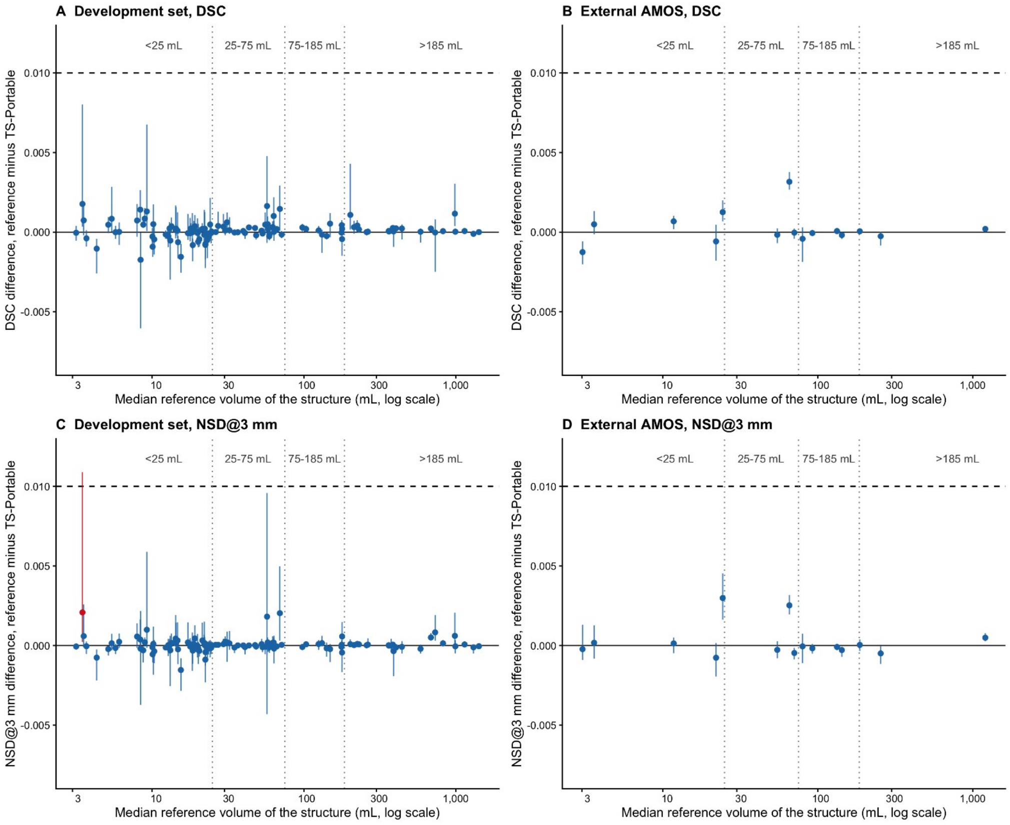

**Supplemental Figure S4.** Man aged 90 or older from the development set. The one structure whose two-sided 95% upper bound exceeded the 0.01 margin: the left 12th rib in the development set (NSD@3 mm difference 0.0021; 95% CI: 0.0002–0.0109; DSC 0.0018; 95% CI: 0.0003–0.0080), driven by the scan shown. Rows show the reference masks of the left 10th (A–C), 11th (D–F), and 12th (G–I) ribs and the labels assigned by the reference implementation and TS-Portable. Colors are the assigned rib label; gray is background. Both implementations assign parts of each reference rib to neighboring rib labels. For the left 12th rib, DSC was 0.181 for the reference implementation, 0.067 for TS-Portable, and 0.188 for the unquantized fp32 models in the same executable. Images are projections of reference voxels colored by assigned label; predictions outside the reference masks are not shown. DSC was computed on the full three-dimensional masks. Magnification is matched within each row. DSC = Dice similarity coefficient; fp32 = 32-bit floating-point.

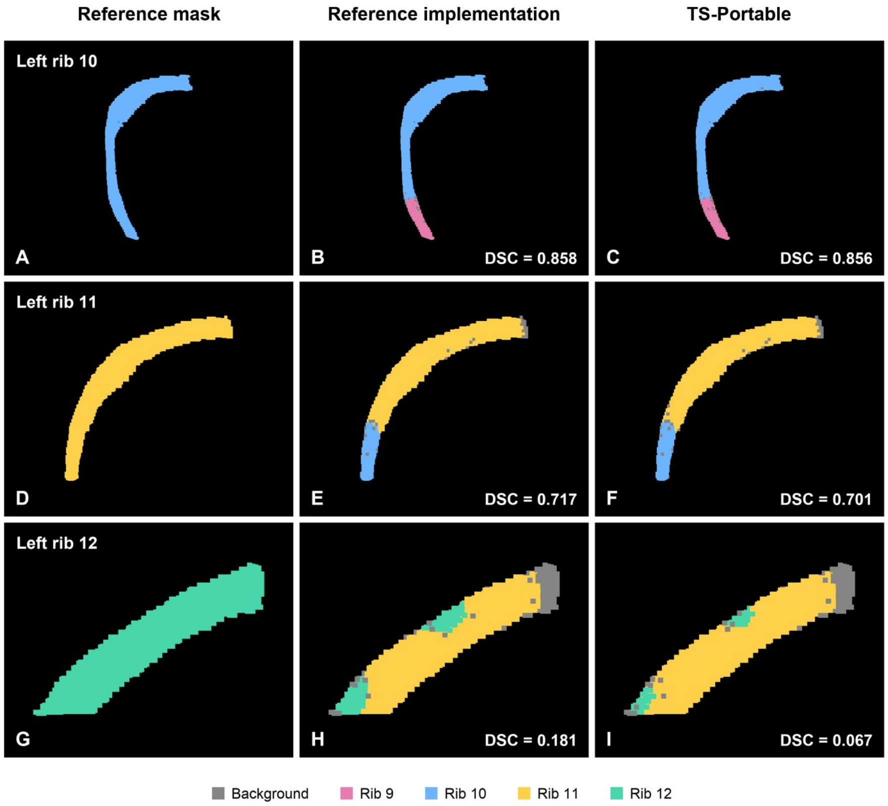

**Supplemental Figure S5.** Woman in her 60s with the smallest spleen in the cohort (16 mL by manual reference), on axial (top) and coronal (bottom) sections through the spleen. (A) Manual spleen (green), the spleen found by the 6-mm localization model (dashed outline; 19 mL), and the spleen-only crop (solid box). (B, C) In spleen-only mode, the reference implementation and TS-Portable labeled 3.4 and 4.9 mL (DSC 0.34 and 0.45). (D) In full 117-label mode, TS-Portable labeled 15 mL (DSC 0.94), as did the reference implementation. The localization model found the spleen and the crop contains all of it; the error arises when the 1.5-mm model segments the crop, which it does in the same way in both implementations. Coronal sections are reformatted from 5-mm axial sections. DSC = Dice similarity coefficient against the manual spleen.

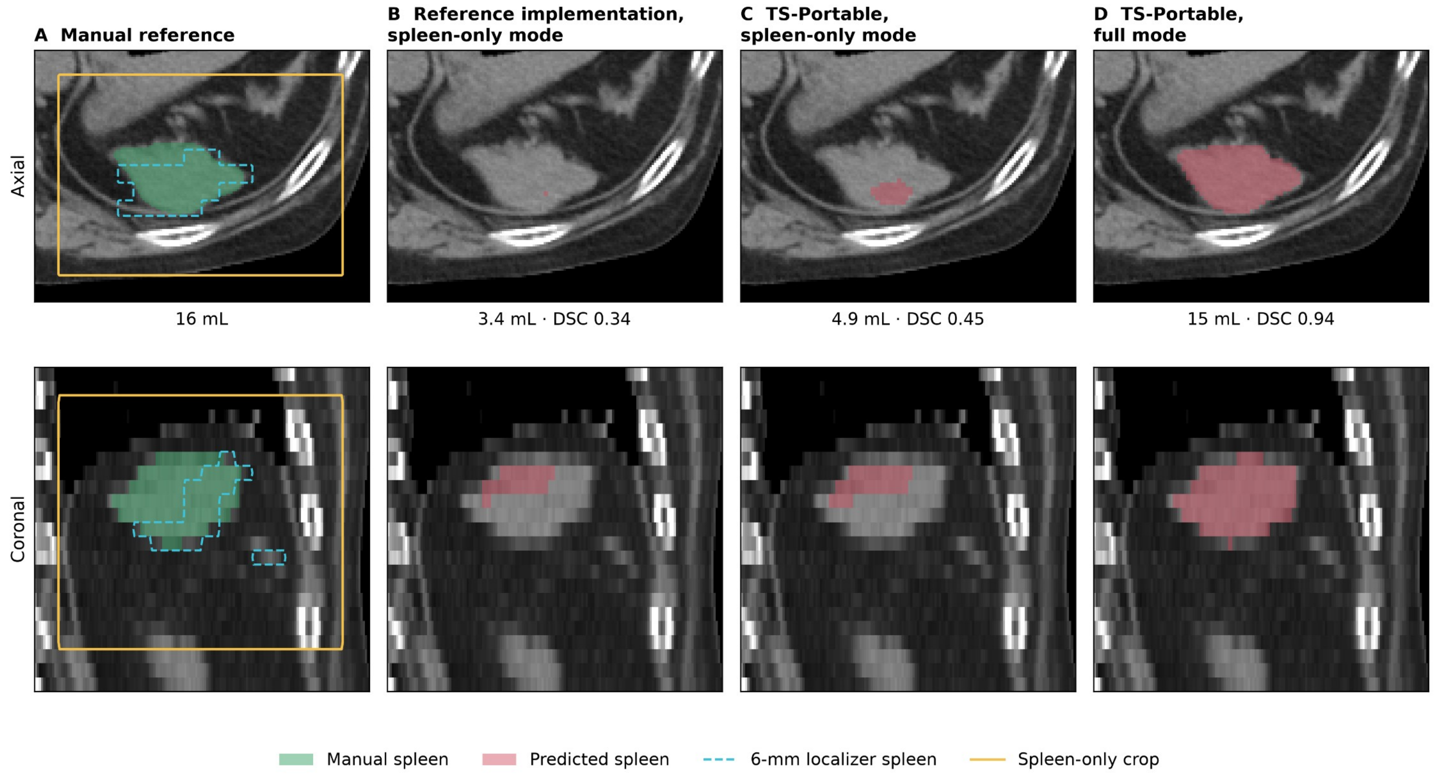
